# Evaluating the CellSearch CMMC Assay for Non-Invasive Longitudinal MRD Monitoring

**DOI:** 10.64898/2026.03.28.26349025

**Authors:** Shemeeakah Powell, Thai Bui, Damodara Gullipalli, Margaret LaCava, Steven Mark Jones, Thomas Hansen, Frank Kuhr, Wojciech Swat, Zoltan Simandi

**Affiliations:** Menarini Silicon Biosystems, Inc.

**Author notes:** Contributed equally (Shemeeakah Powell, Thai Bui, Damodara Gullipalli).

## Abstract

Current clinical management of multiple myeloma (MM) relies on bone marrow (BM) biopsies for minimal residual disease (MRD) assessment. While BM biopsies are the gold standard, their invasive nature and potential to miss extramedullary or patchy disease necessitate sensitive, non-invasive liquid biopsy platforms.

In this study, we evaluated the analytical performance of the CellSearch® CMMC assay to determine its utility for deep-MRD monitoring. Using a standard 4 mL whole blood input, the assay achieves a WBC-normalized sensitivity of 2.45 × 10^−7^, supported by a limit of quantitation of 5 cells per run. Given this high analytical sensitivity, the assay provides a robust negative predictive value, rendering false-negative findings highly unlikely in populations with detectable peripheral disease.

These findings characterize the CellSearch® CMMC assay as a highly sensitive, analytically validated platform for non-invasive deep-MRD level longitudinal surveillance monitoring. When integrated into a clinical workflow that accounts for its specificity profile, the platform offers a patient-friendly complement to serial BM biopsies, with the potential to reduce their frequency in appropriate clinical contexts.

## Introduction

Multiple myeloma (MM) is a genetically complex malignancy defined by the clonal proliferation of malignant plasma cells within the bone marrow (BM) niche. Despite the implementation of transformative therapies, including proteasome inhibitors, immunomodulatory agents, and CAR-T cell modalities, MM typically follows a relapsing-remitting trajectory that requires rigorous, continuous disease monitoring (Swan, Madduri et al. 2024, Alhejazi, Alotaibi et al. 2025). Minimal residual disease (MRD) assessment has become the primary surrogate endpoint for treatment efficacy, with longitudinal MRD negativity serving as a robust predictor of prolonged progression-free and overall survival (Kumar, Paiva et al. 2016, Munshi, Avet-Loiseau et al. 2017, Giri, Dhakal et al. 2025, Landgren and Devlin 2025).

Current clinical standards for MRD detection rely on BM aspiration followed by next-generation flow cytometry (NGF) or next-generation sequencing (NGS), both of which achieve high analytical sensitivity, typically reaching limits of 10^−5^ to 10^−6^ (Kumar, Paiva et al. 2016, Flores-Montero, Sanoja-Flores et al. 2017, Diamond, Rustad et al. 2021, Ding, Xu et al. 2021). However, these methods are constrained by the logistical and biological limitations of BM sampling. Aspirates provide only focal, single-site snapshots of a disease characterized by spatial heterogeneity; furthermore, the invasiveness of repeat procedures and the risk of “dry taps” present significant barriers to the frequent serial monitoring required for real-time tracking of treatment kinetics (Rasche, Chavan et al. 2017).

Liquid biopsy platforms offer a non-invasive, repeatable alternative for systemic disease assessment by interrogating tumor-derived analytes within peripheral blood. Circulating multiple myeloma cells (CMMCs), which traffic from the marrow into systemic circulation, serve as direct targets for these assays (Mishima, Paiva et al. 2017, Gonsalves, Jevremovic et al. 2020). CMMCs provide a critical advantage in cases of non-secretory disease. In non-secretory cases, where “biochemical relapse” is impossible to track, CMMC kinetics may serve as the primary indicator for early intervention. While standard assays for blood proteins, light chains, and electrophoresis often yield false negatives in these patients, CMMC detection remains effective as it identifies the malignant cells directly rather than relying on surrogate protein analytes (Garces, Bretones et al. 2020, Garces, Diamond et al. 2025, Bertamini, Fokkema et al. 2026, Fokkema, Bertamini et al. 2026). The CellSearch® CMMC platform (Menarini Silicon Biosystems) provides an automated solution for this, utilizing immunomagnetic enrichment via CD138-conjugated ferrofluid followed by multi-parameter fluorescent imaging (Foulk, Schaffer et al. 2018).

While CMMC enumeration has historically served as a prognostic indicator, the evolving clinical standard, which now prioritizes the detection of deep minimal residual disease (MRD), demands a rigorous validation of analytical performance to confirm the assay’s ability to reliably identify low-frequency malignant clones at the limits of detection. Given the clinical demand for a high-sensitivity tool to rule out disease progression, understanding the assay’s analytical ceiling and its performance trade-offs is critical.

In this study, we provide a systematic evaluation of the CellSearch® CMMC assay’s performance metrics, including Limit of Blank (LoB), Limit of Quantitation (LoQ), linearity, and inter-instrument reproducibility. By quantifying the assay’s sensitivity against its specificity profile, we define the platform’s utility as a high-confidence tool for deep-MRD monitoring, establishing its potential to provide a patient-centric, non-invasive alternative to serial BM biopsies.

## Materials and Methods

### Ethics and compliance statement

This study was conducted in accordance with the Declaration of Helsinki and all applicable federal regulations governing research involving human subjects. Patient peripheral blood samples were procured through two independent commercial biobanks operating under their own institutional ethical oversight: Analytical Biological Services, Inc. (ABS; Exton, PA, USA), under IRB-approved biobank protocols 313965, 311817, and 313063; and Precision Biospecimens (Louisville, KY, USA), under project protocol 064-ONC-26. Informed consent for the collection, storage, and research use of samples was obtained from all donors by the respective biobanks prior to sample provision. Healthy adult donor recruitment was conducted by Menarini Silicon Biosystems, Inc. under institutional protocol VRXHV-02, approved by Advarra Inc,.. Informed consent was obtained from all healthy donors prior to participation. All samples were de-identified prior to receipt and analysis at Menarini Silicon Biosystems, Inc., and no patient-identifying information was used in any analysis reported herein. All data presented are either aggregated or de-identified in accordance with applicable privacy regulations.

### Patient and healthy donor cohorts

All clinical analyses using patient and healthy donor samples were conducted on peripheral blood collected between 2021 and 2025. Linearity and reproducibility studies were conducted across an extended period spanning July 2018 to September 2025, as detailed in the Proportional Linearity section below. The patient cohort comprised 161 individuals with plasma cell disorders. Patients were stratified into three clinical subgroups based on available clinical metadata: treatment-naïve (n = 96), encompassing newly diagnosed and previously untreated individuals with confirmed plasma cell disorders; treated or in remission (n = 29), including patients who had received at least one prior line of therapy or were in documented clinical remission at the time of collection; and indeterminate treatment status (n = 36), defined as cases for which treatment history could not be ascertained from available biobank records. Healthy adult donors (n = 94) were recruited with no documented prior history of plasma cell dyscrasia, hematological malignancy, or active immunological disease. Donors were not prospectively screened for monoclonal gammopathy by serum protein electrophoresis or serum free light chain analysis. Healthy donor designation therefore represents a potential source of undetected low-level pre-malignant disease, particularly in older donors. Donors spanned a broad age range (18-70+ years) and multiple ethnicities to maximize generalizability of the LoB estimate.

All blood samples were collected into CellRescue® Preservative tubes and processed according to the CellSearch® CMMC standard operating procedure within 120 hours of collection. The 120-hour processing window is consistent with CellRescue® Preservative tube stability specifications. White blood cell (WBC) counts were obtained by Sysmex hematology analyzer at the time of sample processing.

### CellSearch® CMMC assay

The CellSearch® CMMC assay was performed on the CellTracks® AutoPrep® system (Menarini Silicon Biosystems) and analyzed using the CellTracks Analyzer II® semi-automated fluorescence microscope. Peripheral blood (4 mL) was processed by immunomagnetic enrichment targeting the CD138 antigen to capture circulating plasma cells, including circulating multiple myeloma cells (CMMC). Following enrichment, cells were fluorescently labelled with anti-CD38-PE to identify plasma cells, a mixture of anti-CD45-APC and anti-CD19-APC to differentiate leukocytes and B cells, and DAPI for nuclear identification. Enriched and stained cells were transferred to a MagNest® cartridge, which applies a magnetic field to distribute labelled cells across the imaging surface for scanning. Candidate CMMC were scored as CD38⁺, DAPI⁺, CD45⁻, CD19⁻ events by trained operators. All CMMC counts are expressed as cells per 4 mL blood input.

### Generation of the H929-mAzamiGreen (H929-mAG) reporter cell line

NCI-H929 multiple myeloma cells were engineered to express monomeric AzamiGreen (mAzamiGreen) using a CRISPR/Cas9-mediated knock-in approach targeting the AAVS1 safe-harbor locus. The donor homology-directed repair (HDR) template consisted of an EF-1α promoter driving the mAzamiGreen expression cassette, followed by a bovine growth hormone (bGH) polyadenylation signal, flanked by AAVS1-specific homology arms. Ribonucleoprotein (RNP) complexes were assembled by combining recombinant Cas9 nuclease with synthetic guide RNA targeting the AAVS1 locus. These RNP complexes were co-delivered with the HDR donor template into H929 cells via electroporation. Following recovery in standard culture medium, cells expressing mAzamiGreen were enriched via fluorescence-activated cell sorting (FACS). To isolate stable clonal populations, cells were sorted at single-cell density into 96-well plates. Expanded clones were then screened by PCR and Sanger sequencing to confirm targeted genomic integration and stable mAzamiGreen expression. Verified clones were further expanded to establish the H929-mAzamiGreen knock-in cell line.

### H929-mAzamiGreen (H929-mAG) spike-in

For limit of quantitation (LoQ) studies, defined numbers of H929-mAG cells were spiked into background samples using a single-cell dispensing workflow. Cells were first prepared as a single-cell suspension and subjected to fluorescence-based gating using the Pala single-cell dispenser (Bio-Techne), to identify viable mAzamiGreen-positive cells. Live cells were discriminated based on forward/side scatter characteristics and exclusion of debris, followed by selection of mAzamiGreen-positive events. The gated population was then bulk-sorted on the Pala instrument to generate a purified stock of fluorescent, viable cells. Precise numbers of H929-mAG cells were dispensed into recipient samples to create defined spike-in levels across the LoQ range.

### Limit of Blank (LoB), Limit of Detection (LoD) and Limit of Quantitation (LoQ)

The LoB was defined as the 95th percentile of the individual replicate observation counts (n=94), representing the upper boundary of physiologically normal plasma cell circulation in healthy individuals.

The formal analytical LoD, the lowest input concentration at which the probability of detecting at least one cell exceeds 95%, was estimated from these data using the Poisson capture model with an empirical capture rate of λ = 0.50 per spiked cell. Assuming an average recovery rate (λ) of 0.5, the probability of zero-cell recovery is given by e^(−λn). Setting the probability of detecting at least one cell to ≥95% (i.e., 1 − e^(−0.5n) ≥ 0.95) yields a minimum input requirement of approximately 6 cells. Importantly, LoD is reported in units of spiked input cells, while the LoB and LoQ are reported in units of recovered plasma cell counts. At 50% capture efficiency, a spiked input of 6 cells yields an expected recovered count of approximately 3 cells, while the LoQ of 5 reported cells corresponds to a spiked input of approximately 10 cells.

LoQ is most often derived by identifying the lowest analyte concentration at which replicate measurements satisfy a pre-specified precision criterion, typically CV ≤ 20%. Because healthy peripheral blood contains a physiological population of circulating polyclonal plasma cells that the CellSearch® CMMC assay captures indistinguishably from malignant CMMCs, any reported count must first be evaluated against the healthy donor background before it can be interpreted as evidence of disease. The LoQ is therefore governed by the LoB rather than by spike-in precision data alone. Accordingly, the LoQ was set at 5 cells/sample, one integer above the LoB of 4 cells/sample.

### Sensitivity and WBC-normalized detection limit

Analytical sensitivity was evaluated using a genetically engineered H929 cell line stably expressing mAzamiGreen fluorescent protein (H929-mAG). H929-mAG cells were spiked into peripheral blood from three healthy donors at four levels (0, 5, 10, and 50 cells per sample), with 15 replicate measurements per spike level (5 replicates per donor × 3 donors), yielding 60 total measurements. Analysis was restricted to mAzamiGreen-positive events to isolate spiked cells from any background normal plasma cell.

### Proportional linearity

Proportional linearity was evaluated across eight independent studies conducted between July 2018 and September 2025 on the CellSearch® CMMC assay. H929 cells were spiked into healthy donor peripheral blood at four target concentration levels per study (approximately 22, 88, 350, and 1,400 cells/sample), plus a zero-spike negative control. Each study comprised three independent runs, with five replicate measurements per spike level per run, yielding up to 15 non-zero data points per study.

Linearity was assessed by ordinary least-squares (OLS) regression of actual recovered count against target count. Per-study R² values, slopes, and percentage recovery (actual/target × 100) were calculated for all non-zero data points. Acceptable linearity was defined as 80-120% recovery. The grand mean recovery and standard deviation were calculated across all 96 non-zero observations pooled from all eight studies.

### Intra-run precision

Of the 161 patient samples analyzed, 128 had independently processed duplicate plasma cell measurements with the CellSearch® CMMC workflow and were included in the intra-run precision analysis. For each replicate pair, the within-run coefficient of variation was calculated as |rep1 − rep2| / (mean × √2) × 100, providing a bias-corrected estimate of single-measurement dispersion. Replicate pairs were stratified into three categories: both-nonzero (n = 100), both-zero (n = 13, CV = 0% by definition), and one-zero (n = 15, one replicate zero and one non-zero). Precision profiles were examined across five mean-count bins: 0-4, 5-19, 20-99, 100-499, and ≥500 cells/sample. Bland-Altman analysis on log_2_-transformed counts was performed for the 100 both-nonzero pairs to assess systematic bias and limits of agreement (LoA = bias ± 1.96 × SD of log_2_ ratio).

### Inter-instrument reproducibility

Inter-instrument reproducibility was evaluated in two independent concordance studies. In Study 2025a, seven samples were each run in duplicate on two instruments. In Study 2025b, the same seven samples were run in duplicate, with one instrument serving as the common comparator part of both studies. For each sample and instrument, the two replicate counts were averaged prior to pairwise comparison. Inter-instrument agreement was assessed by paired Wilcoxon signed-rank test, Bland-Altman analysis of count differences, and visual inspection of paired dot plots. One replicate from Study 2025a (Instrument B; raw count 44 vs. paired replicate 26) was identified as a statistical outlier and excluded from aggregate LoA calculations.

## Results

### Healthy donor background and Limit of Blank (LoB) threshold determination

The CellSearch® CMMC kit enables enrichment and enumeration of circulating plasma cells in peripheral blood (**Figure 1A**). However, low levels of circulating plasma cells can be present in the peripheral blood of healthy individuals as part of normal immune homeostasis. This physiological background must be characterized and distinguished from disease-associated circulating malignant plasma cells. To establish the assay’s biological specificity, plasma cell counts were measured in 94 healthy donors with no known hematological malignancy. Demographic characteristics including age, gender, and ethnicity are summarized in **Supplemental Table 1** and **Supplemental Figure 1**. The distribution of plasma cell counts was markedly right-skewed, with the majority of donors yielding zero or low cell counts (**Figure 1B**). Specifically, 55.3% of donors (n = 52) returned a plasma cell count of exactly zero, 39.4% (n = 37) yielded between one and four cells per run, and 5.3% (n = 5) returned five or more cells per run. The highest count observed in any healthy donor was 12 cells per 4 mL blood. Based on this distribution, the LoB was established at 4 cells per sample, corresponding to the 95th percentile of the healthy donor population (**Figure 1B**). To confirm that healthy donor measurements remained consistently below this threshold while also assessing repeatability across independent runs, samples from 20 donors were each analyzed in eight replicate experiments (n = 160 total observations; **Figure 1C**). Nineteen of twenty donors (95%) remained below the LoB across all eight replicates. A single donor produced one replicate reaching the LoB (maximum observed count = 4 cells/sample). Per-donor mean counts ranged from 0 to 2.25 cells per sample, confirming that the healthy donor background is stable and reproducibly under the above established biological specificity range (**Figure 1C**).

**Figure 1.**
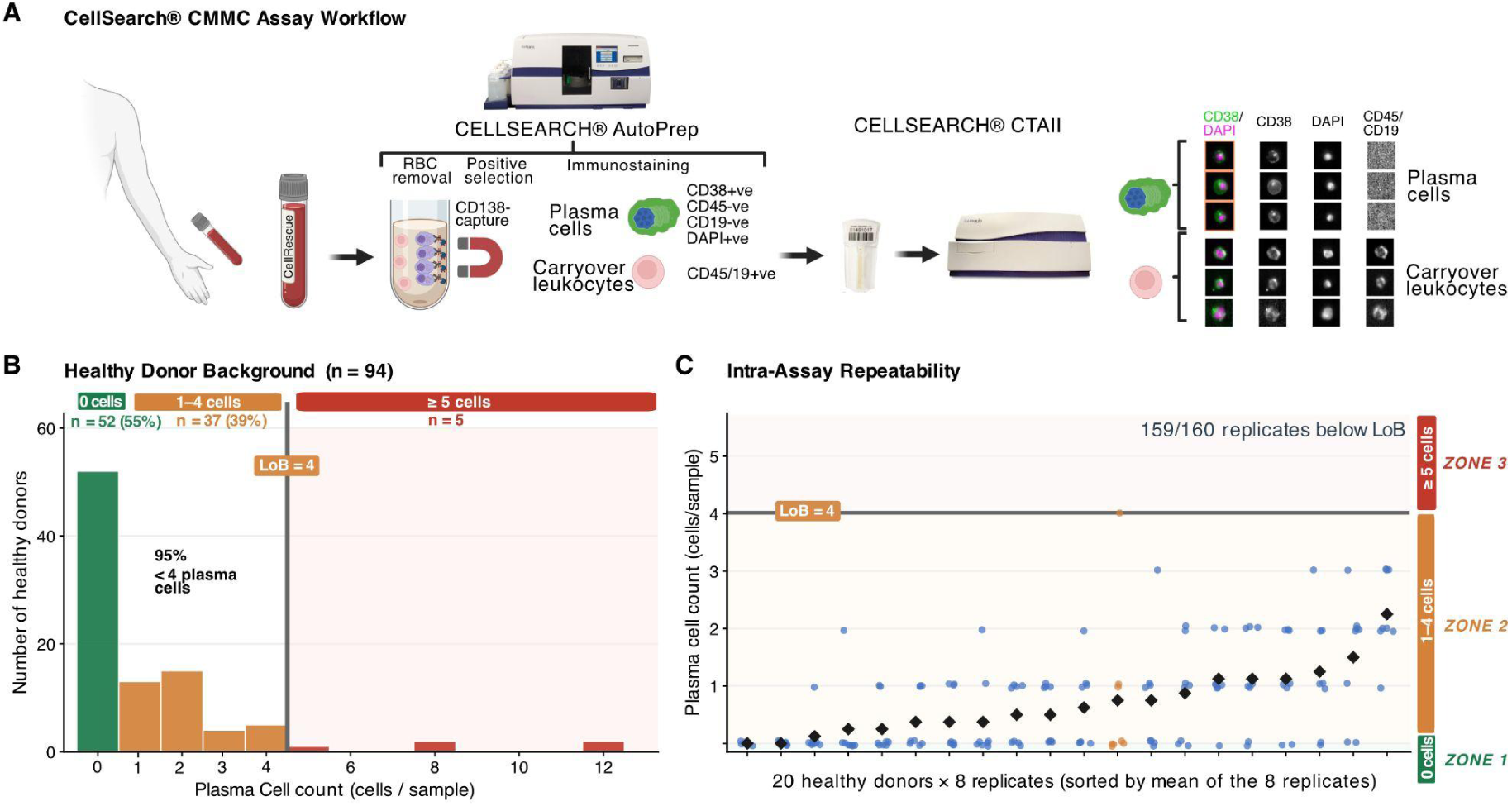
Establishment of the biologically defined Limit of Blank for the CellSearch® CMMC assay using healthy donor samples. **(A)** Schematic overview of the CellSearch® CMMC assay workflow. Peripheral blood is collected into CellRescue® preservative tubes and subjected to immunomagnetic enrichment using anti-CD138, followed by fluorescent labelling (anti-CD38-PE; anti-CD45/CD19-APC; DAPI), automated scanning on the CellTracks Analyzer II, and plasma cell enumeration. Results are reported as cells per 4 mL run. **(B)** Distribution of plasma cell counts across 94 healthy donors. Bars are colored according to a three-zone classification: Zone 1 (plasma cell count = 0, green; n = 52, 55.3%), Zone 2 (1-4 cells, orange; n = 37, 39.4%), and Zone 3 (≥ 5 cells, red; n = 5, 5.3%). **(C)** Intra-assay repeatability assessed in 20 healthy donors each measured across 8 independent replicates (n = 160 observations total). Individual replicate counts are shown as jittered circles colored by whether the donor ever reached the LoB (orange, n = 1) or remained consistently below it (blue, n = 19). Black diamonds indicate per-donor mean counts. **Abbreviations:** CMMC, circulating multiple myeloma cell; LoB, limit of blank; PE, phycoerythrin; APC, allophycocyanin; DAPI, 4′,6-diamidino-2-phenylindole.

Importantly, the established 4 cells/sample LoB does not reflect analytical noise from the instrument or reagents, but rather defines the upper limit of physiologically normal plasma cell circulation in healthy individuals.

### Characterization of the analytical Limit of Detection (LoD)

While the LoB sets a biological specificity threshold, the analytical Limit of Detection (LoD), the lowest number of cells that the platform can reliably distinguish from a true zero signal, must be determined independently using a detection channel free of biological background. To accomplish this, a spike-in study was conducted using H929-mAG cells, a genetically engineered derivative of the NCI-H929 myeloma cell line that constitutively expresses the mAzamiGreen (mAG) fluorescent protein from the AAVS1 safe-harbor locus (**Figure 2A**). Since no endogenous cells in healthy peripheral blood emit mAzamiGreen fluorescence, this model provides a true zero-background detection channel orthogonal to the immunophenotypic CMMC panel.

**Figure 2.**
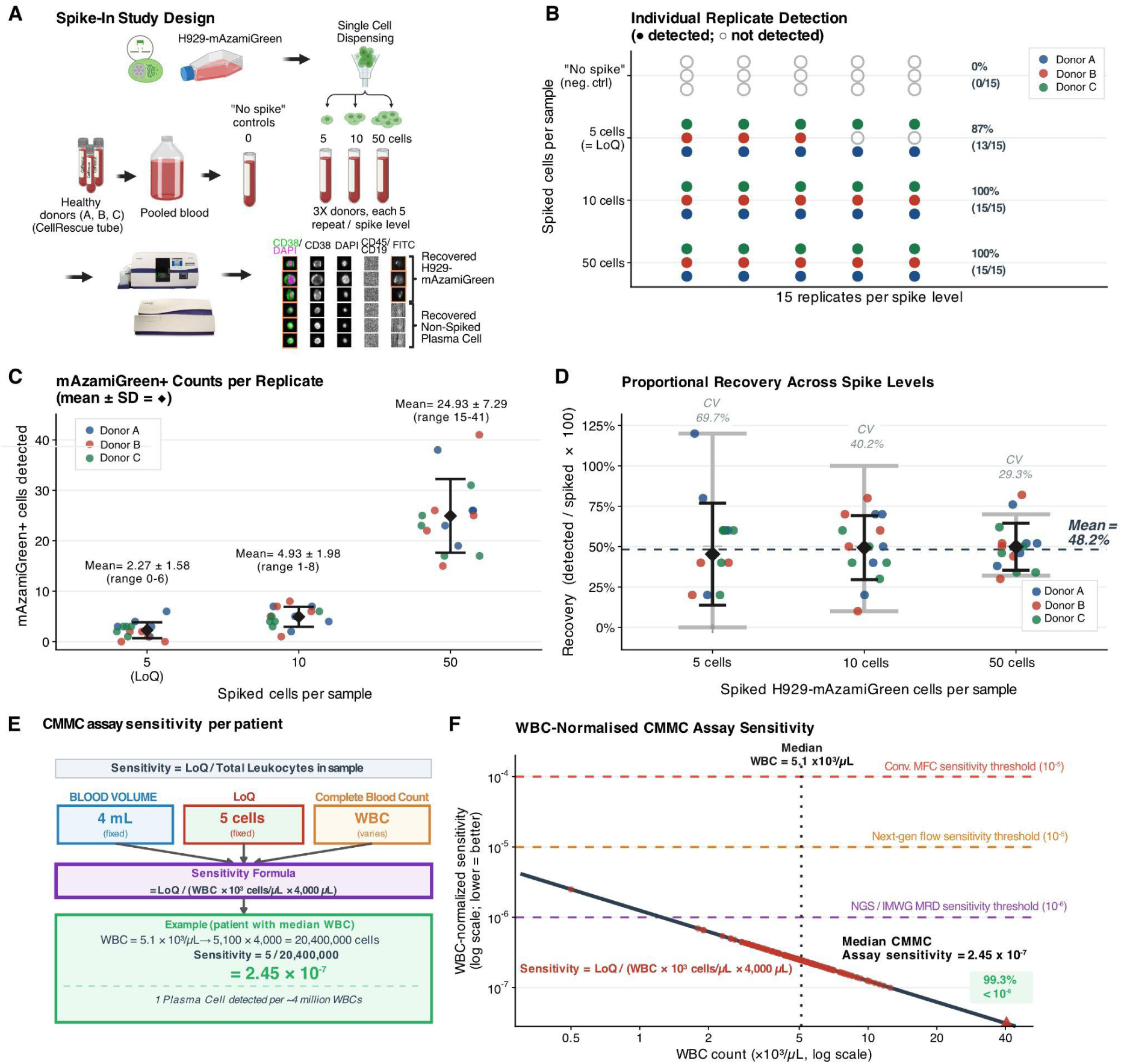
mAzamiGreen spike-in study establishing LoD, LoQ, and capture efficiency. **(A)** Schematic of the spike-in study design. Genetically engineered H929 cells stably expressing the mAzamiGreen fluorescent protein (H929-mAG) were spiked into peripheral blood from three healthy donors (Donor A, B and C) at four concentration levels: 0 (“No spike” control), 5, 10, and 50 cells per 4 mL blood. Five independent replicates were performed per donor per level, yielding n = 15 replicates at each spike level and n = 60 samples in total. **(B)** Per-replicate detection matrix. Each circle represents one replicate; filled donor-colored circles indicate H929-mAG detection, open grey circles indicate non-detection. Detection rates are shown (n detected / n total). **(C)** mAzamiGreen-positive counts per individual replicate at spike levels 5, 10, and 50 cells/sample. Each point represents one replicate, colored by the donor. Black diamonds indicate per-level means; error bars indicate ± 1 SD. **(D)** Proportional recovery by spike level. Y-axis shows recovery expressed as a percentage (mAzamiGreen-positive cells detected / cells spiked × 100) for each of the n = 15 replicates at each level. Donor-colored filled circles indicate detected replicates. Black diamonds and error bars show mean ± 1 SD per level. The horizontal dashed line marks the overall mean recovery of 48.2% across all levels. Grey whiskers indicate the exact Poisson 95% prediction interval for the expected number of captured cells given a capture rate of λ ≈ 0.50 per spiked cell. Coefficient of variation (CV) per level is annotated inside each column. **(E)** Schematic illustrating the derivation of per-patient WBC-normalized analytical sensitivity. Three quantities determine CMMC assay sensitivity: the fixed blood input volume (4 mL = 4,000 µL), the assay LoQ (5 cells/sample), and the sample-specific WBC count from a concurrent complete blood count (CBC). The sensitivity formula is: Sensitivity = LoQ / (WBC × 10³ cells/µL × 4,000 µL). At the cohort median WBC (5.1 × 10³ cells/µL) a sample yields a sensitivity of 2.45 × 10^−7^. **(F)** WBC-normalized sensitivity plotted against WBC count (both axes log_10_ scale) for n = 146 MM patients with available CBC data. Each patient’s value lies on the theoretical hyperbola Sensitivity = 5 / (WBC × 4 × 10^6^), shown as a dark regression line. Horizontal dashed lines indicate published MRD detection methods’s sensitivity as reference: conventional multiparameter flow cytometry (conv. MFC, 10^−4^, red), next-generation flow cytometry (10^−5^, orange), and the IMWG NGS threshold (10^−6^, purple). The vertical dotted line marks the cohort median WBC (5.1 × 10³ cells/µL). The blue vertical band indicates the normal WBC reference range (4.5-11.0 × 10³ cells/µL). One patient (ID ABS311817-08; WBC = 40.4 × 10³ cells/µL) with a markedly elevated WBC is highlighted separately. Median sensitivity = 2.45 × 10^−7^. **Abbreviations**: mAG, mAzamiGreen; CV, coefficient of variation; LoQ, limit of quantitation; SD, standard deviation; WBC, white blood cell count; MRD, minimal residual disease; MFC, multiparameter flow cytometry; NGS, next-generation sequencing; IMWG, International Myeloma Working Group;

H929-mAG cells were spiked into 4 mL whole peripheral blood from three healthy donors at four target concentrations (0, 5, 10, and 50 cells per sample), with five independent replicates per donor per level (n = 15 per level; n = 60 total samples) (**Figure 2A**). In all 15 negative-control “No spike” replicates, zero mAzamiGreen-positive events were recorded, confirming the complete absence of background signal in the orthogonal fluorescence channel (**Figure 2B**). At 5 spiked cells per sample, 13 of 15 replicates yielded at least one mAzamiGreen-positive cell event, corresponding to a detection rate of 86.7%. At 10 and 50 spiked cells per sample, detection was 100% across all 15 replicates at each level. mAzamiGreen-positive counts per replicate increased proportionally with the number of spiked cells across all three donors (**Figure 2C**). Mean observed counts were 2.27 ± 1.58 at 5 cells per sample, 4.93 ± 1.98 at 10 cells per sample, and 24.93 ± 7.29 at 50 cells per sample. Recovery, expressed as the ratio of detected to spiked cells, was 45.3% (SD 31.6%) at 5 cells per sample, 49.3% (SD 19.8%) at 10 cells per sample, and 49.9% (SD 14.6%) at 50 cells per sample. These data yield an overall mean capture efficiency of 48.2%, corresponding to an empirical capture rate of λ ≈ 0.50 per spiked cell (**Figure 2D**).

Using this empirically derived capture rate, variability across replicates was evaluated in the context of Poisson counting statistics. The coefficient of variation decreased with increasing spike concentration, from 69.7% at 5 cells per sample to 40.2% at 10 cells per sample and 29.3% at 50 cells per sample, consistent with the expected inverse square root relationship between relative dispersion and event count (**Figure 2D**).

Based on this model, the analytical LoD was estimated using a Poisson framework with λ = 0.50 per spiked cell. The LoD is defined as the lowest input concentration at which the probability of detecting at least one cell exceeds 95%, and is therefore expressed in units of spiked input cells, distinct from the LoB and LoQ, which are expressed in units of recovered (reported) counts. Accordingly, a minimum input of approximately 6 spiked cells is required to achieve ≥95% detection probability, consistent with the observed transition from 86.7% detection at 5 spiked cells to 100% at 10 spiked cells. At the empirically derived capture efficiency of ∼50%, an input of 6 spiked cells yields an expected recovered count of approximately 3 cells. This is well below the LoQ of 5 recovered cells, confirming that the assay can physically detect cells at concentrations below the biological specificity boundary.

### Defining the Limit of Quantitation Based on biological specificity

Having established that the CellSearch® CMMC assay can physically detect cells at concentrations below the biological specificity boundary, a separate and clinically critical question is at what reported cell count can a positive result be confidently asserted to exceed the range of normal plasma cell physiology rather than reflect physiological background trafficking? In case of the CellSearch® CMMC assay this question cannot be answered by the analytical LoD alone, since the LoD is derived against a true zero background and carries no information about where normal plasma cell counts end and pathological CMMC burden begins. The Limit of Quantitation (LoQ) was therefore determined to define the lowest reported cell count that can be assigned a clinically meaningful positive interpretation, the threshold above which a result can be reported with quantifiable confidence as exceeding the healthy donor plasma cell distribution. Accordingly, the Limit of Quantitation (LoQ) was set at 5 cells per sample (reported in units of recovered counts), one cell above the biologically defined LoB of 4 cells per sample, ensuring that reported counts exceed the normal physiological background (**Figure 2E**). This represents the lowest reportable count that exceeds the 95th percentile of the healthy donor plasma cell distribution, such that a reported count of 5 or more cells per sample carries a less than 5% probability of arising from normal physiological plasma cell trafficking.

### WBC-normalized analytical sensitivity

Because the CellSearch® CMMC assay enumerates cells from a fixed blood volume (4 mL), the effective sensitivity relative to total circulating leukocytes is not a fixed property of the assay but varies as a function of each patient’s WBC count at the time of blood collection. To make this relationship explicit, we derived a per-patient sensitivity metric according to the formula: Sensitivity = LoQ / (WBC × 10³ cells/µL × 4,000 µL), where WBC is expressed in units of 10³ cells/µL as reported by a standard complete blood count (CBC) (**Figure 2E**). Under this formulation, all variability in sensitivity across patients is attributable exclusively to inter-individual differences in WBC count; the LoQ and blood volume are assay constants. A total of 161 patients with plasma cell disorders, including newly diagnosed (NDMM), relapsed/refractory (RRMM), and remission/MRD-negative cases, were enrolled. Sensitivity analysis was performed in the subset of 146 patients for whom concurrent complete blood count (CBC) data were available (**Figure 2F**). Patient WBC counts ranged from 0.5 to 40.4 × 10³ cells/µL (median 5.1, IQR 3.9-6.2 × 10³ cells/µL), consistent with the range expected in a treated myeloma population. One patient (ABS311817-08) presented with a markedly elevated WBC count of 40.4 × 10³ cells/µL, likely reflecting concurrent leucocytosis, but did not affect summary statistics. Across the cohort, WBC-normalized sensitivity ranged from 3.09 × 10^−8^ to 2.50 × 10^−6^, with a median of 2.45 × 10^−7^. While BM-based and blood-based assays measure fundamentally different biological fractions and cannot be directly compared for clinical sensitivity, to contextualise the scale of the CellSearch® CMMC assay blood-based sensitivity, the WBC-normalized values are shown alongside the typical detection thresholds established for bone marrow-based MRD platforms (conventional multiparameter flow cytometry (10^−4^), next-generation flow cytometry (10^−5^), and the IMWG-endorsed NGS threshold of 10^−6^)) (**Figure 2F**).

100% of patients achieved sensitivity better than the conventional multiparameter flow cytometry threshold of 10^−4^, the next-generation flow cytometry threshold of 10^−5^, and 99.3% (145/146) reached sensitivity below the International Myeloma Working Group (IMWG)-endorsed NGS threshold of 10^−6^ (Kumar, Paiva et al. 2016, Leonardos, Benetatos et al. 2025). While some emerging platforms report analytical sensitivities exceeding these typical referenced levels, the CMMC assay overall demonstrates robust performance against established methods.

Taken together, these results establish that the CellSearch® CMMC assay achieves sub-10^−6^ WBC-normalized sensitivity in essentially all patients presenting with a normal circulating leukocyte count, positioning it as analytically competitive with next-generation molecular blood compartment MRD platforms.

### Verification of analytical linearity across instruments and operators

To evaluate the accuracy of the CellSearch® CMMC assay across its dynamic range, proportional linearity was assessed using samples with known expected CMMC counts. Proportional linearity of the CellSearch® CMMC assay was evaluated across eight independent studies, including multiple reagent lots, operators, and instrument systems. Each study followed a common design in which samples with known expected CMMC counts were prepared and analyzed, with the resulting actual counts compared to the expected input. Across all 96 pooled observations, regression of actual versus expected counts yielded an Ordinary Least Squares (OLS) slope of 1.00 with an intercept of 4.2 cells per sample and an overall R^2^ of 0.991, demonstrating near-perfect proportionality across the entire concentration range (**Figure 3A**). On a per-study basis, the R^2^ ranged from 0.981 to 0.999, and per-study slopes ranged from 0.994 to 1.004, indicating that no individual study introduced a systematic bias in either direction. The consistency of slopes across studies separated by up to seven years is particularly notable given the involvement of different reagent lots and different instruments.

**Figure 3.**
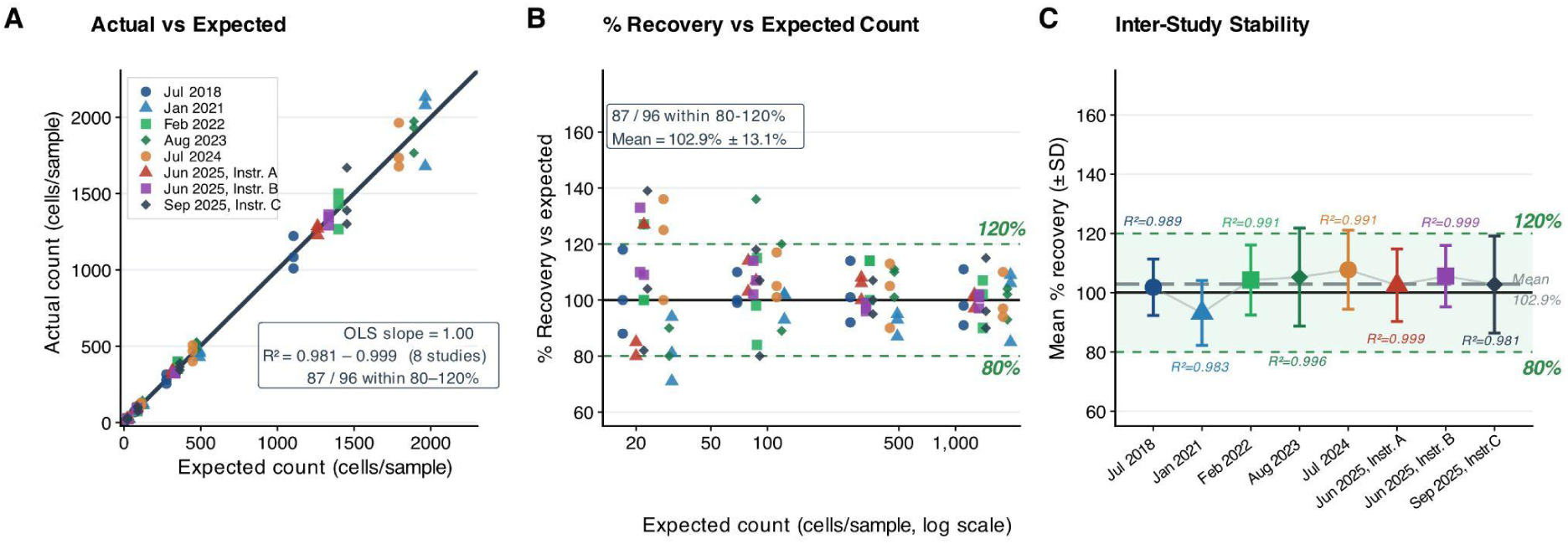
Proportional linearity of the CellSearch® CMMC assay. **(A)** Actual versus expected cell counts for all 96 data points pooled across eight linearity studies. Each point represents one measurement at a defined expected concentration; colors and shapes identify individual studies. The dashed grey line is the identity (y = x, perfect recovery). The solid dark line is the pooled ordinary least-squares (OLS) regression fit. The inset box reports the OLS slope (1.00), the per-study R^2^ range (0.981-0.999), and the total proportion of data points falling within the 80-120% recovery acceptance range (87/96). **(B)** Percentage recovery relative to the expected count, plotted on a log_10_-scaled x-axis. Each point represents the percentage recovery for one measurement (actual / expected × 100). The green-shaded dashed lines mark acceptance boundaries; the solid black line indicates 100% recovery. The inset reports the total number of points within the acceptance range and the grand mean recovery ± SD. **(C)** Per-study mean percentage recovery (± 1 SD) plotted across the eight studies. Individual study means are shown as colored symbols; error bars indicate ± 1 SD. The dashed horizontal green lines mark the 80-120% acceptance range; the solid black line indicates 100% recovery; the long-dashed grey line indicates the mean across all studies. The per-study R^2^ value is annotated for each time point. Studies conducted on three separately designated instruments (Instr. A, B and C), demonstrating cross-instrument reproducibility. **Abbreviations:** OLS, ordinary least squares; SD, standard deviation.

Across all 96 measurements, 87 (90.6%) fell within the pre-specified 80-120% acceptance range (**Figure 3B**). The mean recovery was 102.9% (SD 13.1%, median 101.5%), indicating negligible systematic bias relative to the expected input (**Figure 3C**). The full recovery range spanned 71% to 139%; the nine out-of-range observations comprised eight exceeding 120% and one falling below 80%. When the distribution of out-of-range points is examined by expected concentration, they are disproportionately concentrated at the lowest end of the tested range, where Poisson counting variability produces inherently wider relative dispersion. Above approximately 70 cells per sample, all measurements fell within the acceptance range.

Per-study mean recoveries showed no directional trend (**Figure 3C**), with the mean of 102.9% remaining stable across all time points. The three studies, run concurrently on three instruments, returned mean recoveries of 102.5% (SD 12.2%), 105.6% (SD 10.4%), and 102.8% (SD 16.4%), with per-study R^2^ values of 0.999, 0.999, and 0.981, demonstrating that the linearity performance of the assay is reproducible across independent instrument platforms operated in parallel.

Taken together, these data establish that the CellSearch® CMMC assay is linear across a clinically relevant dynamic range, free from systematic count-dependent bias, and consistent across time and multiple instruments..

### Evaluation of intra-run precision

Following confirmation of proportional assay performance across the tested concentration range, analytical precision was evaluated using clinical samples. Intra-run precision was assessed in a cohort of 128 MM patient samples, each processed in duplicate through the complete CellSearch® CMMC assay workflow (**Figure 4A**). For each replicate pair, the coefficient of variation was calculated as |rep1 -rep2| / (mean × √2) × 100, providing a bias-corrected, single-measurement estimate of within-run dispersion. The 128 replicate pairs were stratified into three subsets: (i) 100 pairs with non-zero counts in both replicates, enabling CV estimation; (ii) 13 pairs with zero counts in both replicates; and (iii) 15 discordant pairs in which one replicate yielded zero and the other a non-zero count.

**Figure 4.**
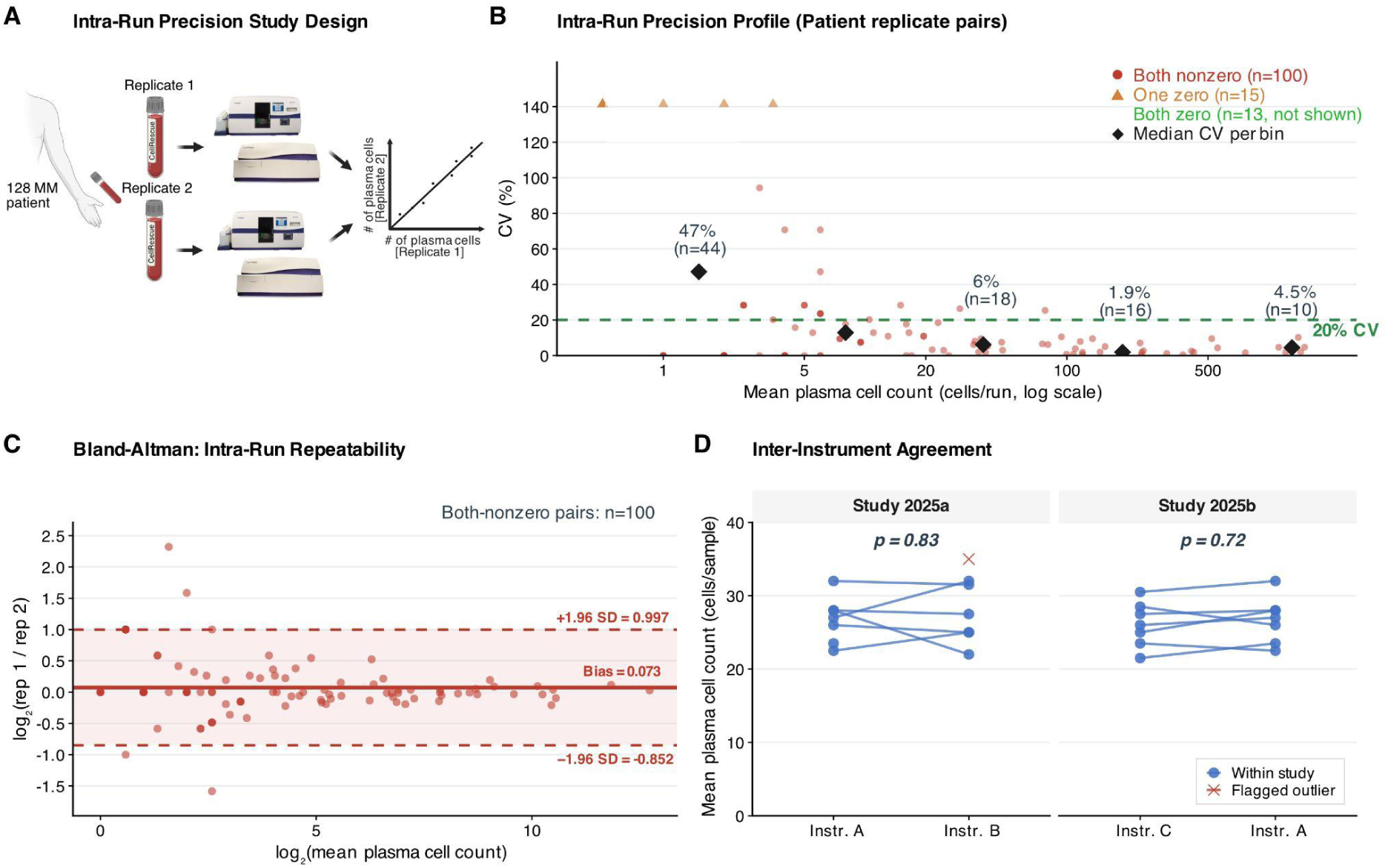
Intra-run precision and inter-instrument reproducibility of the CellSearch® CMMC assay. **(A)** Schematic overview of the intra-run precision study design. Patient peripheral blood from 128 patients were each processed twice through the full CellSearch® CMMC workflow in independent parallel runs. **(B)** Intra-run precision profile for all 128 patient replicate pairs, plotted as CV (%) against the mean plasma cell count on a log_10_-scaled x-axis. For each of the 128 replicate pairs generated, the coefficient of variation (CV) was calculated as |rep1 − rep2| / (mean × √2) × 100 to provide a bias-corrected estimate of within-run dispersion. Pairs in which both replicates returned zero counts (n = 13) were treated separately and assigned CV = 0% (100% concordant). Pairs in which only one replicate returned zero (n = 15) are flagged as one-zero pairs and are shown in the precision profile. Black diamonds indicate the median CV within each count bin (0-4, 5-19, 20-99, 100-499, ≥500 cells/sample); per-bin medians and sample sizes are annotated. The dashed green line marks the 20% CV threshold. **(C)** Bland-Altman analysis of intra-run repeatability for the 100 both-nonzero replicate pairs. The y-axis shows the log_2_ ratio (rep 1 / rep 2); the x-axis shows log_2_(mean count). The solid red line indicates the mean bias; dashed red lines indicate ±1.96 SD limits of agreement (LoA). Both-zero pairs (n = 13, CV = 0%) are excluded from this plot as their log_2_ ratio is undefined. **(D)** Inter-instrument agreement across two concordance studies (Study 2025a: Instruments A vs. B; Study 2025b: Instruments C vs. A). In each study, 7 patient samples were each measured in duplicate on each of the two instruments; the duplicate counts were averaged before pairing. Each line connects the mean counts for the same sample across the two instruments. A single replicate from Study 2025a was flagged as an outlier and excluded from inter-study comparisons; it is shown as a red × and its connecting line is omitted. P-values are from the study-level paired equivalence test (Study 2025a: p = 0.83; Study 2025b: p = 0.72). **Abbreviations:** CV, coefficient of variation; LoA, limits of agreement; SD, standard deviation.

The precision profile showed the characteristic hyperbolic relationship between CV and plasma cell count magnitude, consistent with Poisson counting statistics. Median CV decreased from 47% in the lowest count bin (mean 0-4 cells/sample; n = 29 pairs) to 6% at 20-99 cells/sample (n = 18) and further to 1.9% and 4.5% in the 100-499 (n = 16) and ≥500 cells/sample (n = 10) bins, respectively (**Figure 4B**). At and above the LoQ of 5 cells/sample, 60 of 71 both-nonzero pairs (85%) achieved CV < 20%. Overall, 77 of 100 both-nonzero pairs (77%) fell below the 20% CV threshold across the dynamic range. The 13 both-zero pairs were 100% concordant (CV = 0%). The 15 one-zero pairs, in which one replicate detected one or more cells while the paired replicate returned zero, represent the expected probabilistic consequence of sampling rare cells near the detection limit and are consistent with the 86.7% detection rate established at the LoD in the spike-in study.

On the log_2_ scale, the mean bias across 100 both-nonzero pairs was 0.073 log_2_ units (equivalent to a 1.05-fold change), confirming the absence of large directional bias between replicate runs (**Figure 4C**). Ninety of 100 pairs (90%) fell within ±1 log_2_ unit (2-fold change). No systematic trend in the ratio with increasing mean count was apparent across the dynamic range of the dataset, indicating that precision does not decrease at higher plasma cell counts.

Inter-instrument reproducibility was evaluated in two concordance studies conducted on three CellSearch® instruments, each testing 7 patient samples in duplicate per instrument. Duplicate measurements on instruments A and B across 7 patient samples were equivalent (paired test p = 0.83) (**Figure 4D**). A single replicate (resulting cell count 44) from instrument B was identified as an outlier and excluded from inter-study analyses. Among the remaining 6 clean pairs, all differences fell within 6 cells/sample. Duplicate measurements on instruments A and C across all 7 patient samples were likewise equivalent (paired test p = 0.72), with no outliers detected (**Figure 4D**).

These findings demonstrate that CellSearch® CMMC assay plasma cell counts are quantitatively equivalent across distinct instruments and timepoints, supporting longitudinal monitoring.

### CMMC count distributions define three reporting zones

While analytical linearity and precision established the assay performance, these are not sufficient to define the assay’s clinical utility. Interpretation of measured counts requires parameters that translate analytical output into clinically meaningful categories. For the CMMC assay, this is governed by two key metrics: the Limit of Quantitation (LoQ), defined on Figure 2, and the negative predictive value (NPV), which reflects the probability that a negative result represents true absence of detectable disease rather than limited assay sensitivity. To avoid confounding effects from treatment-induced cell depletion, only treatment-naïve patients were included in the performance analysis. CMMC counts from treatment-naïve patients with plasma cell disorders (n = 96) and healthy donors (n = 94) were stratified into three reporting zones: *Zone 1* (0 cells per sample; undetectable), *Zone 2* (1–4 cells per sample; below the LoQ and indeterminate), and *Zone 3* (≥5 cells per sample; at or above the LoQ) (**Figure 5A**). These groups showed strong separation across the full dynamic range (Wilcoxon rank-sum, W = 7,645.5, p = 4.5 × 10^−17^).

**Figure 5.**
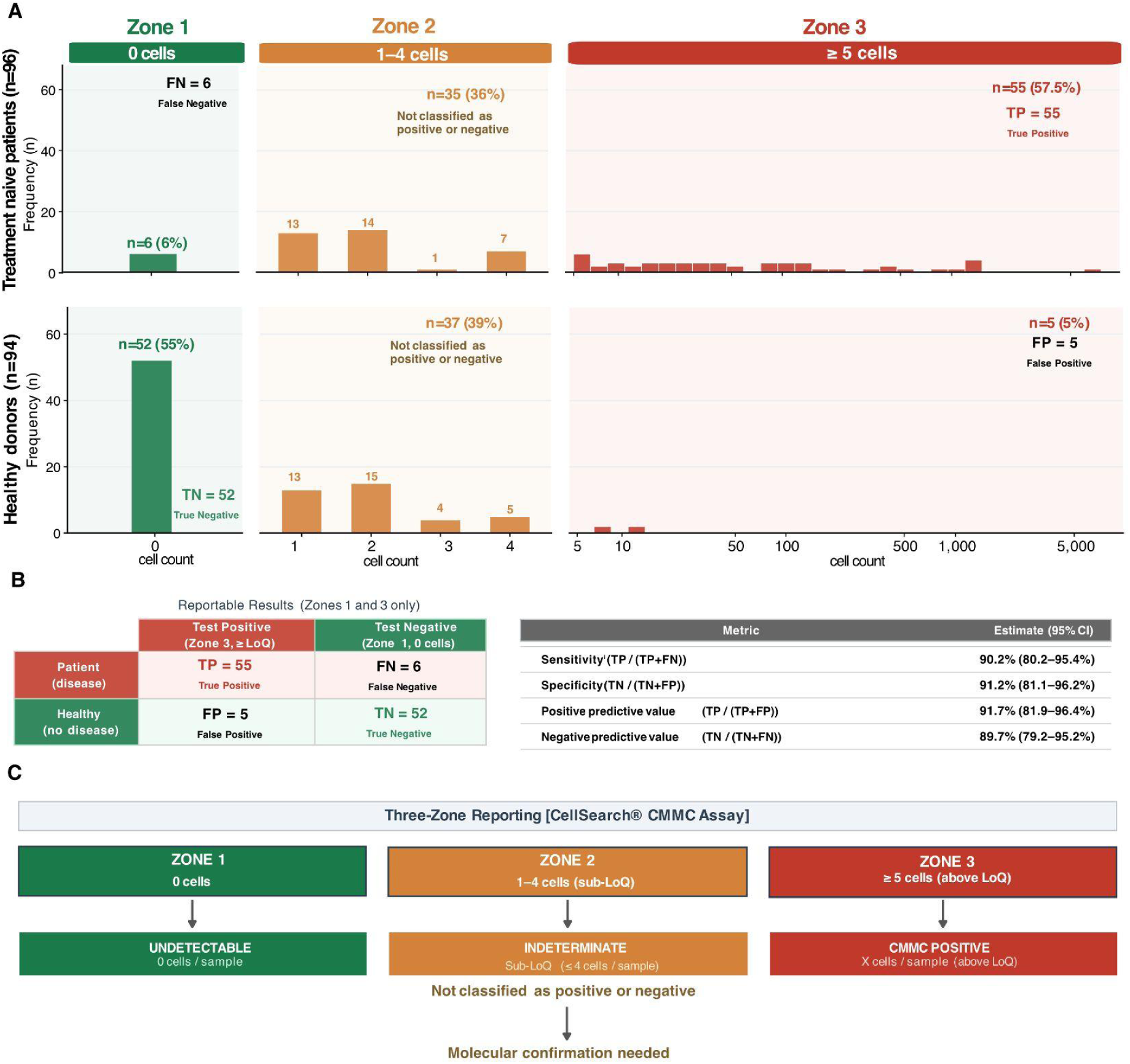
Three-zone framework for interpretation of CellSearch® CMMC assay plasma cell counts. **(A)** Frequency distributions of mean CMMC counts in treatment-naïve patients with plasma cell disorders (top, n = 96) and healthy donors (bottom, n = 94). Counts are displayed in three reporting zones defined by their relationship to the limit of quantitation (LoQ = 5 cells/run): *Zone 1* (green, 0 cells/sample; undetectable), *Zone 2* (orange, 1-4 cells/sample; sub-LoQ, indeterminate), and *Zone 3* (red, ≥ 5 cells/sample; at or above the LoQ, quantifiable). *Zone 3* is rendered as a histogram on a log_10_(plasma cell + 1) axis, with axis labels showing original CMMC values in cells/sample. The y-axis scale is held constant within each cohort row across all three zones. **(B)** Clinical performance of the three-zone classification framework. Left: contingency table for reportable results only (*Zone 1* = negative CMMC report, *Zone 3* = positive CMMC report). *Zone 2* results (1-4 cells/sample) are indeterminate and are excluded from all metric denominators. Right: performance metrics with 95% Wilson score confidence intervals computed over reportable results. Sensitivity reflects the *Zone 3* detection rate among reportable treatment-naïve patients. **(C)** Three-zone classification framework summarizing the clinical decision logic associated with each zone result. **Abbreviations:** TP, true positive; TN, true negative; FP, false positive; FN, false negative.

Among healthy donors, 52 of 94 samples (55.3%) returned zero counts (*Zone 1, True Negative*) and 37 of 94 (39.4%) returned counts in the 1-4 cells/sample range (*Zone 2*), consistent with the low-level background signal attributable to normal circulating plasma cells. Five of 94 healthy donors (5.3%) yielded counts at or above the LoQ (*Zone 3*, *False Positive*), defining an analytical false-positive rate of 5.3% at the quantification threshold. The distribution in treatment-naïve patients was shifted toward higher plasma cell counts. Overall, 57% of patients (55/96) fell within *Zone 3*, with quantifiable CMMC count (median across the cohort: 6 cells per sample; true positives). An additional 36.5% (35/96) had cell counts in *Zone 2* (1–4 cells per sample), while 6.2% (6/96) had no detectable cells (*Zone 1*; false negatives). Counts in the range of 1–4 cells per sample (*Zone 2*) cannot be classified as either positive or negative. This range substantially overlaps with the previously established healthy donor background, with 39.4% of healthy donors falling within this interval, and lies below the LoQ, where individual measurements are not analytically distinguishable from stochastic assay variation. Accordingly, *Zone 2* results are classified as indeterminate and excluded from the denominator of all performance metrics.

Within the treatment-naïve cohort, 35 of 96 patients (36.5%) and 37 of 94 healthy donors (39.4%) fell into the indeterminate category, corresponding to an overall indeterminate rate of 37.9% (72 of 190 samples). This represents a quantifiable trade-off inherent to the zone-based classification framework.

Restricting analysis to the 61 reportable patients and 57 reportable healthy donors (*Zone 1* and *Zone 3* only), the 2×2 contingency table yielded TP = 55, FN = 6, FP = 5, TN = 52 (**Figure 5B**). *Zone 3* sensitivity among reportable results was 90.2% (95% CI 80.2-95.4%) and specificity was 91.2% (95% CI 81.1-96.2%). Positive and negative predictive values at the empirical mix of reportable samples were 91.7% (95% CI 81.9-96.4%) and 89.7% (95% CI 79.2-95.2%), respectively. These metrics characterize the classification performance of the assay within its reportable range and are distinct from the analytical sensitivity of 86.7% established by controlled spike-in experiments (**Figure 2**), which quantifies the probability of detection at the LoQ under optimal conditions rather than the clinical detection rate in patient samples.

Together, these data define a three-zone framework in which each category carries a distinct interpretation: *Zone 1* supports probabilistic rule-out, *Zone 3* provides a quantifiable disease signal suitable for longitudinal monitoring, and *Zone 2* defines a sub-LoQ indeterminate range requiring orthogonal confirmation before clinical decision-making (**Figure 5C**).

## Discussion

Multiple myeloma management is entering an era defined by depth of response. The progressive adoption of quadruplet induction regimens, novel immunotherapy combinations, and maintenance strategies has shifted the clinical question from whether a remission is achieved to whether residual disease is truly absent at the molecular level (Engelhardt, Kortum et al. 2024, Szalat, Anderson et al. 2024). In this context, the value of any MRD monitoring platform is determined less by its ability to detect active disease and more by its confidence in a negative result. The present study establishes the analytical performance characteristics of the CellSearch® CMMC assay across a comprehensive set of validation parameters, including sensitivity, linearity, precision, inter-instrument reproducibility, and specificity, and demonstrates that the resulting NPV profile places it firmly within the range of clinical utility for longitudinal liquid biopsy monitoring.

The most consequential finding of this validation is the WBC-normalized sensitivity of 2.45 × 10^−7^, achieved from a standard 4 mL blood draw. At this sensitivity, a single CMMC among approximately four million white blood cells is detectable, a level of resolution that approaches the theoretical minimum imposed by Poisson sampling statistics at the input blood volume used.

The specificity of 94.7% (89/94 healthy donors below the LoQ) is analytically valid and reflects the biological reality of circulating plasma cell physiology. A non-trivial proportion of healthy individuals harbor low numbers of plasma cells in the peripheral blood, as has been documented by multiple groups independent of the CellSearch® platform (Caraux, Klein et al. 2010). In this cohort, 39.4% of healthy donors showed between 1 and 4 cells per sample (counts detectable by the assay but falling below the LoQ threshold) and 5.3% returned counts at or above 5 cells per sample. The three-zone labeling was designed explicitly to accommodate this biology. *Zone 2* (1-4 cells/sample) results are designated indeterminate, acknowledging the sub-LoQ range as uninterpretable. Critically, the false-positive events in the healthy cohort were bounded in magnitude. The maximum healthy donor count was 12 cells/sample, which would typically fall well below the quantitative CMMC counts seen in active, untreated disease (median >100 cells per sample in newly diagnosed patients (Foulk, Schaffer et al. 2018), though overlap with treated or deep-remission patients near the detection boundary warrants clinical awareness. Of note, because Monoclonal Gammopathy of Undetermined Significance (MGUS) is often asymptomatic and prevalent in older populations (occurring in approximately 3% of individuals over age 50), a presumed healthy donor pool may inadvertently include individuals with low-level circulating pre-myeloma cells representing a potentially confounding aspect in the establishment of LoB/LoD of the assay.

The most tractable approach to resolving this specificity constraint is orthogonal molecular confirmation of the immunophenotypically enriched cells. The CellSearch® workflow is uniquely positioned for this purpose. Following fluorescence imaging and enumeration, the enriched cellular material remains physically present on the cartridge in a fixed, immunostained state. This downstream accessibility enables the same sample that generated the CMMC count to serve as input material for subsequent molecular characterization without additional blood draw or sample processing. For patients in the *Zone 2* or low *Zone 3* range, where the distinction between normal plasma cell trafficking and residual malignant disease cannot be resolved by morphology and immunophenotype alone, molecular confirmation of clonality provides a direct resolution path.

The most rigorous approach to clonality confirmation is IGH V(D)J rearrangement sequencing of the enriched CMMC fraction (Martinez-Lopez, Lahuerta et al. 2014, Oberle, Brandt et al. 2017, Rustad, Hultcrantz et al. 2019, Hultcrantz, Rustad et al. 2022). Each malignant plasma cell clone carries a unique immunoglobulin heavy-chain rearrangement established at the time of B-cell commitment; normal, polyclonal plasma cells in the peripheral blood of healthy individuals will present as a diverse mixture of rearrangements. A CMMC sample in which a single dominant V(D)J clonotype accounts for a substantial fraction of sequencing reads provides strong evidence of malignant monoclonal origin, regardless of cell count magnitude. In the context of a *Zone 2* indeterminate result or a low-positive *Zone 3* result from a patient in suspected remission, this clonality determination directly answers the diagnostic question that count magnitude alone cannot.

Beyond clonality confirmation, the post-imaging CMMC fraction is a tractable substrate for the full spectrum of somatic molecular characterization relevant to myeloma biology and clinical management. Single-nucleotide variant (SNV) analysis of established myeloma driver genes, including those in the RAS/MAPK pathway (KRAS, NRAS, BRAF), the NF-κB pathway (TRAF3, CYLD, BIRC2/3), TP53, and FAM46C, can be performed on CMMC-derived DNA using targeted amplicon or capture-based sequencing panels with sufficient sensitivity for low-input material (Barwick, Gupta et al. 2019, Alberge, Dutta et al. 2025, Garces, Diamond et al. 2025, Fokkema, Bertamini et al. 2026). Detection of structural variants including the high-risk translocations t(4;14), t(14;16), and t(11;14), as well as del(17p), del(1p), and gain(1q), can be accomplished either by the FISH approach described in earlier CMMC studies (Foulk, Schaffer et al. 2018), or increasingly by sequencing of the enriched fraction, which simultaneously reports copy number across the entire genome and has the practical advantage of not requiring pre-selection of target loci.

Protein-level characterization with the CellSearch® CMMC assay is equally accessible. Immunophenotypic markers of therapeutic resistance, notably reduced expression of CD38 (the target of daratumumab and isatuximab), can be assessed on the enriched fraction using dedicated image analysis software. In the context of the accelerating deployment of CD38-directed therapies in both newly diagnosed and relapsed/refractory disease, real-time immunophenotypic monitoring of target antigen expression directly from blood represents a clinically meaningful capability that bone marrow biopsy provides only intermittently and at procedural cost to the patient.

The growing recognition of blood-based MRD assessment as a clinically valid monitoring strategy provides an important regulatory and clinical framework for the CellSearch® CMMC assay. The International Myeloma Working Group (IMWG) MRD consensus criteria, alongside emerging guidance from regulatory agencies, increasingly acknowledge that non-invasive MRD assessment modalities, including circulating tumor DNA (ctDNA) and circulating tumor cell enumeration, can provide complementary information or, in specific contexts, serve as surrogate endpoints to bone marrow-based methods, particularly in the longitudinal monitoring setting (Thakral, Das et al. 2020). The practical advantages are substantial. Blood draws are well-tolerated, repeatable at high frequency without patient burden, and not subject to the spatial sampling limitations inherent in focal or patchy marrow disease, where a single-site biopsy may underestimate or entirely miss extramedullary plasmacytoma or heterogeneous marrow involvement. The present study demonstrates that the CellSearch® CMMC assay is a high-confidence rule-out tool that can supplement, and under appropriate clinical circumstances reduce the frequency of, serial bone marrow biopsies.

It is worth distinguishing the CellSearch® CMMC assay from the ctDNA-based liquid biopsy approaches with which it is increasingly compared. Cell-free circulating tumor DNA reflects the aggregate mutational landscape of the tumor but requires knowledge of the patient-specific mutation or IGH clonotype at baseline to enable tracking, and its sensitivity is highly dependent on tumor burden and shedding rate (Mithraprabhu, Khong et al. 2017, Thakral, Das et al. 2020). The CMMC approach captures intact tumor cells that can be independently enumerated, phenotyped, and molecularly characterized without prior knowledge of a target sequence. In practice, the two modalities are complementary. ctDNA provides quantitative sensitivity at deep MRD levels driven by high mutational burden, while CMMC provides cellular identity confirmation and direct molecular characterization capability at the single-cell or low number of pooled cell levels. A combinatorial workflow in which both assays are applied to the same blood draw would provide both quantitative depth and qualitative cellular resolution across the full dynamic range of myeloma disease monitoring.

Several limitations of the current study need to be acknowledged. The healthy donor specificity cohort (n = 94), while sufficient for analytical validation, does not comprehensively sample the clinical populations most likely to generate borderline results, including patients with monoclonal gammopathy of undetermined significance (MGUS), smouldering multiple myeloma in deep imaging response, or post-CAR-T patients with incomplete immune reconstitution. Expansion of the specificity characterization to include these populations would refine the *Zone 2* and low *Zone 3* interpretation boundaries and provide disease-specific false-positive rates more directly applicable to clinical decision-making.

A key limitation of the CellSearch® CMMC assay is measurement uncertainty at low cell counts due to Poisson sampling. Intra-run variability showed 11.7% discordance between replicate pairs (one zero, one non-zero), making *Zone 1* (0 cells) and *Zone 2* (1–4 cells) or *Zone 3* results difficult to distinguish at a single time point near the LoQ. However, this limitation is mitigated in longitudinal monitoring, where clinical interpretation relies on trends across serial measurements rather than individual results. Consistent patterns over time provide more reliable evidence of disease status, and clinical decisions should be based on multi-draw trajectories rather than single measurements.

A critical distinction that the present validation study is not designed to resolve is the relationship between the assay’s analytical sensitivity and its clinical sensitivity against bone marrow MRD status. Trafficking of malignant plasma cells from the marrow niche into peripheral blood is biologically variable and not linearly proportional to marrow burden, particularly in patients receiving effective therapy. Resolving this relationship requires a prospective concordance study in which same-day paired CMMC enumeration and BM-based NGF or NGS MRD assessment are obtained across defined treatment timepoints, with sufficient representation of patients at MRD thresholds of 10^−5^ and 10^−6^ to characterize the detection concordance rate as a function of marrow burden. Importantly, however, even in the absence of formal concordance data, the practical asymmetry between the two sampling modalities has independent clinical value. Bone marrow aspiration is limited to infrequent, procedure-intensive timepoints, making it unsuitable for capturing treatment kinetics in real time. In contrast, the blood-based CMMC assay enables serial sampling aligned with clinical decision cycles, supporting longitudinal trajectory analysis, a clinically orthogonal dimension of disease monitoring that a single concordance snapshot cannot provide.

In summary, the CellSearch® CMMC assay, as analytically characterized in this study, represents a technically mature, quantitative, and molecularly extensible platform for the liquid biopsy monitoring of multiple myeloma. Its defining analytical strength lies in a WBC-normalized sensitivity of 2.45 × 10^−7^ from a standard blood draw, supported by proportional linearity across the measured range and inter-instrument agreement comparable to within-instrument variability, establishing a robust quantitative foundation for longitudinal CMMC monitoring. The resulting NPV profile, anchored by the three-zone classification framework developed here, provides clinically actionable negative predictive confidence across the prevalence range of greatest clinical interest. The 5.3% *Zone-3* false-positive rate, the primary specificity constraint of the platform, is addressable through downstream molecular characterization of the enriched cellular fraction, an approach uniquely enabled by the CellSearch® architecture and not available to cell-free ctDNA platforms. Integrated with clonality sequencing, targeted SNV analysis, and structural variant profiling, the CellSearch® CMMC assay has the potential to deliver, from a single non-invasive blood draw, the combination of quantitative tumor burden, phenotypic identity, and molecular disease characterization that currently requires serial, invasive bone marrow procedures.

## Data Availability

All data produced in the present study are available upon request to the authors

## Author Contributions

S.P., T.B., and D.G. contributed equally to this work. S.P., T.B., D.G., M.L., and S.M.J. performed experiments and analyzed data. T.H., F.K., and W.S. conceived and designed the study. Z.S. conceived and designed the study, analyzed data, created visualizations, performed statistical analysis and wrote the manuscript. All authors reviewed and approved the final manuscript.

## Data Availability Statement

Aggregate and de-identified data underlying the figures and tables in this manuscript are provided in full in the Supplementary Materials. Proprietary assay configuration parameters are commercially confidential and are not available for disclosure.

## References

Alberge, J. B., A. K. Dutta, A. Poletti, T. H. H. Coorens, E. D. Lightbody, R. Toenges, X. Loinaz, S. Wallin, A. Dunford, O. Priebe, J. Dagan, C. J. Boehner, E. Horowitz, N. K. Su, H. Barr, L. Hevenor, K. Towle, R. Beesam, J. B. Beckwith, J. Perry, D. M. Cordas Dos Santos, L. Bertamini, P. T. Greipp, K. Kubler, P. F. Arndt, C. Terragna, E. Zamagni, E. M. Boyle, K. Yong, G. Morgan, B. A. Walker, M. A. Dimopoulos, E. Kastritis, J. Hess, R. Sklavenitis-Pistofidis, C. Stewart, G. Getz and I. M. Ghobrial (2025). “Genomic landscape of multiple myeloma and its precursor conditions.” Nat Genet 57(6): 1493–1503.

Alhejazi, A., G. S. Alotaibi, A. S. A. Saleh, M. Alahmadi, I. Motabi, F. Z. Alsharif, A. Alamer, O. Abduljalil, I. Tailor, M. Marei, A. S. Barefah, M. Aljabry, S. Alhayli, B. Usman, A. Hanbali, A. Alabdulwahab, I. ElHemaidi, H. M. Alahwal, E. Mutahar and A. Alsaeed (2025). “Guidelines on Management of Multiple Myeloma in the Relapsed/Refractory Setting: The Saudi Myeloma Working Group Guideline.” Clin Lymphoma Myeloma Leuk 25(10): e756–e765.

Barwick, B. G., V. A. Gupta, P. M. Vertino and L. H. Boise (2019). “Cell of Origin and Genetic Alterations in the Pathogenesis of Multiple Myeloma.” Front Immunol 10: 1121.

Bertamini, L., C. Fokkema, P. Rodriguez-Otero, M. van Duin, E. Terpos, M. D’Agostino, V. H. J. van der Velden, N. van de Donk, M. Delforge, C. Driessen, R. Hajek, H. Einsele, A. Vangsted, D. Vieyra, R. Attar, A. Sitthi-Amorn, R. Carson, F. Schjesvold, P. Robak, M. Beksac, A. Spencer, A. Broijl, T. Cupedo, P. Moreau, M. Boccadoro and P. Sonneveld (2026). “Circulating tumor cells predict myeloma outcomes in patients treated with daratumumab, bortezomib, lenalidomide, and dexamethasone.” Blood 147(4): 431–442.

Caraux, A., B. Klein, B. Paiva, C. Bret, A. Schmitz, G. M. Fuhler, N. A. Bos, H. E. Johnsen, A. Orfao, M. Perez-Andres and N. Myeloma Stem Cell (2010). “Circulating human B and plasma cells. Age-associated changes in counts and detailed characterization of circulating normal CD138- and CD138+ plasma cells.” Haematologica 95(6): 1016–1020.

Diamond, B. T., E. Rustad, K. Maclachlan, K. Thoren, C. Ho, M. Roshal, G. A. Ulaner and C. O. Landgren (2021). “Defining the undetectable: The current landscape of minimal residual disease assessment in multiple myeloma and goals for future clarity.” Blood Rev 46: 100732.

Ding, H., J. Xu, Z. Lin, J. Huang, F. Wang, Y. Yang, Y. Cui, H. Luo, Y. Gao, X. Zhai, W. Pang, L. Zhang and Y. Zheng (2021). “Minimal residual disease in multiple myeloma: current status.” Biomark Res 9(1): 75.

Engelhardt, M., K. M. Kortum, H. Goldschmidt and M. Merz (2024). “Functional cure and long-term survival in multiple myeloma: how to challenge the previously impossible.” Haematologica 109(8): 2420–2435.

Flores-Montero, J., L. Sanoja-Flores, B. Paiva, N. Puig, O. Garcia-Sanchez, S. Bottcher, V. H. J. van der Velden, J. J. Perez-Moran, M. B. Vidriales, R. Garcia-Sanz, C. Jimenez, M. Gonzalez, J. Martinez-Lopez, A. Corral-Mateos, G. E. Grigore, R. Fluxa, R. Pontes, J. Caetano, L. Sedek, M. C. Del Canizo, J. Blade, J. J. Lahuerta, C. Aguilar, A. Barez, A. Garcia-Mateo, J. Labrador, P. Leoz, C. Aguilera-Sanz, J. San-Miguel, M. V. Mateos, B. Durie, J. J. M. van Dongen and A. Orfao (2017). “Next Generation Flow for highly sensitive and standardized detection of minimal residual disease in multiple myeloma.” Leukemia 31(10): 2094–2103.

Fokkema, C., L. Bertamini, M. M. E. de Jong, S. Tahri, D. Hofste Op Bruinink, Z. Kellermayer, N. Papazian, C. den Hollander, M. P. W. Vermeulen, E. C. G. Stoetman, G. van Beek, R. Hoogenboezem, V. H. J. van der Velden, C. Hulin, A. Perrot, P. Moreau, M. Rowe, D. Vieyra, R. Carson, M. van Duin, M. A. Sanders, A. Broijl, P. Sonneveld and T. Cupedo (2026). “Circulating tumor cells in myeloma are a compound biomarker for bone marrow high-risk genomic alterations and tumor load.” Blood 147(9): 932–945.

Foulk, B., M. Schaffer, S. Gross, C. Rao, D. Smirnov, M. C. Connelly, S. Chaturvedi, M. Reddy, G. Brittingham, M. Mata, M. Repollet, C. Rojas, D. Auclair, M. DeRome, M. C. Network, B. Weiss and A. K. Sasser (2018). “Enumeration and characterization of circulating multiple myeloma cells in patients with plasma cell disorders.” Br J Haematol 180(1): 71–81.

Garces, J. J., G. Bretones, L. Burgos, R. Valdes-Mas, N. Puig, M. T. Cedena, D. Alignani, I. Rodriguez, D. A. Puente, M. G. Alvarez, I. Goicoechea, S. Rodriguez, M. J. Calasanz, X. Agirre, J. Flores-Montero, L. Sanoja-Flores, P. Rodriguez-Otero, R. Rios, J. Martinez-Lopez, P. Millacoy, L. Palomera, R. Del Orbe, A. Perez-Montana, H. El Omri, F. Prosper, M. V. Mateos, L. Rosinol, J. Blade, J. J. Lahuerta, A. Orfao, C. Lopez-Otin, J. F. San Miguel, B. Paiva and G. P. c. s. group (2020). “Circulating tumor cells for comprehensive and multiregional non-invasive genetic characterization of multiple myeloma.“ Leukemia 34(11): 3007–3018.

Garces, J. J., B. Diamond, T. Sevcikova, S. Nenarokov, D. Bilek, E. Radova, O. Venglar, V. Kapustova, R. Firestone, K. Maclachlan, A. Simhal, L. Broskevicova, J. Vrana, L. Muronova, T. Popkova, J. Mihalyova, H. Plonkova, M. Durante, B. Ziccheddu, M. Simicek, H. J. Cho, G. Mulligan, J. Keats, D. Zihala, O. Landgren, R. Hajek, S. Usmani, F. Maura and T. Jelinek (2025). “Elevated circulating tumor cells reflect high proliferation and genomic complexity in multiple myeloma.” Hemasphere 9(9): e70218.

Giri, S., B. Dhakal, N. S. Callander, E. Medvedova, K. Godby, B. R. Dholaria, S. Bal, G. Ravi, S. Chhabra, R. W. Silbermann and L. Costa (2025). “Optimal MRD-based end point to support response-adapted treatment cessation in newly diagnosed multiple myeloma.” Blood 146(6): 707–716.

Gonsalves, W. I., D. Jevremovic, B. Nandakumar, A. Dispenzieri, F. K. Buadi, D. Dingli, M. Q. Lacy, S. R. Hayman, P. Kapoor, N. Leung, A. Fonder, M. Hobbs, Y. L. Hwa, E. Muchtar, R. Warsame, T. V. Kourelis, S. Russell, J. A. Lust, Y. Lin, R. S. Go, M. A. Siddiqui, R. A. Kyle, M. A. Gertz, S. V. Rajkumar and S. K. Kumar (2020). “Enhancing the R-ISS classification of newly diagnosed multiple myeloma by quantifying circulating clonal plasma cells.” Am J Hematol 95(3): 310–315.

Hultcrantz, M., E. H. Rustad, V. Yellapantula, A. Jacob, T. Akhlaghi, N. Korde, S. Mailankody, A. M. Lesokhin, H. Hassoun, E. L. Smith, O. B. Lahoud, H. J. Landau, G. L. Shah, M. Scordo, D. J. Chung, S. Giralt, E. Papaemmanuil and O. Landgren (2022). “Capture Rate of V(D)J Sequencing for Minimal Residual Disease Detection in Multiple Myeloma.” Clin Cancer Res 28(10): 2160–2166.

Kumar, S., B. Paiva, K. C. Anderson, B. Durie, O. Landgren, P. Moreau, N. Munshi, S. Lonial, J. Blade, M. V. Mateos, M. Dimopoulos, E. Kastritis, M. Boccadoro, R. Orlowski, H. Goldschmidt, A. Spencer, J. Hou, W. J. Chng, S. Z. Usmani, E. Zamagni, K. Shimizu, S. Jagannath, H. E. Johnsen, E. Terpos, A. Reiman, R. A. Kyle, P. Sonneveld, P. G. Richardson, P. McCarthy, H. Ludwig, W. Chen, M. Cavo, J. L. Harousseau, S. Lentzsch, J. Hillengass, A. Palumbo, A. Orfao, S. V. Rajkumar, J. S. Miguel and H. Avet-Loiseau (2016). “International Myeloma Working Group consensus criteria for response and minimal residual disease assessment in multiple myeloma.” Lancet Oncol 17(8): e328–e346.

Landgren, O. and S. M. Devlin (2025). “Minimal Residual Disease as an Early Endpoint for Accelerated Drug Approval in Myeloma: A Roadmap.” Blood Cancer Discov 6(1): 13–22.

Leonardos, D., L. Benetatos, E. Apostolidou, E. Koumpis, L. Dova, E. Kapsali, I. Kotsianidis and E. Hatzimichael (2025). “Applications of Multiparameter Flow Cytometry in the Diagnosis, Prognosis, and Monitoring of Multiple Myeloma Patients.” Diseases 13(10).

Martinez-Lopez, J., J. J. Lahuerta, F. Pepin, M. Gonzalez, S. Barrio, R. Ayala, N. Puig, M. A. Montalban, B. Paiva, L. Weng, C. Jimenez, M. Sopena, M. Moorhead, T. Cedena, I. Rapado, M. V. Mateos, L. Rosinol, A. Oriol, M. J. Blanchard, R. Martinez, J. Blade, J. San Miguel, M. Faham and R. Garcia-Sanz (2014). “Prognostic value of deep sequencing method for minimal residual disease detection in multiple myeloma.” Blood 123(20): 3073–3079.

Mishima, Y., B. Paiva, J. Shi, J. Park, S. Manier, S. Takagi, M. Massoud, A. Perilla-Glen, Y. Aljawai, D. Huynh, A. M. Roccaro, A. Sacco, M. Capelletti, A. Detappe, D. Alignani, K. C. Anderson, N. C. Munshi, F. Prosper, J. G. Lohr, G. Ha, S. S. Freeman, E. M. Van Allen, V. A. Adalsteinsson, F. Michor, J. F. San Miguel and I. M. Ghobrial (2017). “The Mutational Landscape of Circulating Tumor Cells in Multiple Myeloma.” Cell Rep 19(1): 218–224.

Mithraprabhu, S., T. Khong, M. Ramachandran, A. Chow, D. Klarica, L. Mai, S. Walsh, D. Broemeling, A. Marziali, M. Wiggin, J. Hocking, A. Kalff, B. Durie and A. Spencer (2017). “Circulating tumour DNA analysis demonstrates spatial mutational heterogeneity that coincides with disease relapse in myeloma.” Leukemia 31(8): 1695–1705.

Munshi, N. C., H. Avet-Loiseau, A. C. Rawstron, R. G. Owen, J. A. Child, A. Thakurta, P. Sherrington, M. K. Samur, A. Georgieva, K. C. Anderson and W. M. Gregory (2017). “Association of Minimal Residual Disease With Superior Survival Outcomes in Patients With Multiple Myeloma: A Meta-analysis.” JAMA Oncol 3(1): 28–35.

Oberle, A., A. Brandt, M. Voigtlaender, B. Thiele, J. Radloff, A. Schulenkorf, M. Alawi, N. Akyuz, M. Marz, C. T. Ford, A. Krohn-Grimberghe and M. Binder (2017). “Monitoring multiple myeloma by next-generation sequencing of V(D)J rearrangements from circulating myeloma cells and cell-free myeloma DNA.” Haematologica 102(6): 1105–1111.

Rasche, L., S. S. Chavan, O. W. Stephens, P. H. Patel, R. Tytarenko, C. Ashby, M. Bauer, C. Stein, S. Deshpande, C. Wardell, T. Buzder, G. Molnar, M. Zangari, F. van Rhee, S. Thanendrarajan, C. Schinke, J. Epstein, F. E. Davies, B. A. Walker, T. Meissner, B. Barlogie, G. J. Morgan and N. Weinhold (2017). “Spatial genomic heterogeneity in multiple myeloma revealed by multi-region sequencing.” Nat Commun 8(1): 268.

Rustad, E. H., M. Hultcrantz, V. D. Yellapantula, T. Akhlaghi, C. Ho, M. E. Arcila, M. Roshal, A. Patel, D. Chen, S. M. Devlin, A. Jacobsen, Y. Huang, J. E. Miller, E. Papaemmanuil and O. Landgren (2019). “Baseline identification of clonal V(D)J sequences for DNA-based minimal residual disease detection in multiple myeloma.” PLoS One 14(3): e0211600.

Swan, D., D. Madduri and J. Hocking (2024). “CAR-T cell therapy in Multiple Myeloma: current status and future challenges.” Blood Cancer J 14(1): 206.

Szalat, R., K. Anderson and N. Munshi (2024). “Role of minimal residual disease assessment in multiple myeloma.” Haematologica 109(7): 2049–2059.

Thakral, D., N. Das, A. Basnal and R. Gupta (2020). “Cell-free DNA for genomic profiling and minimal residual disease monitoring in Myeloma-are we there yet?” Am J Blood Res 10(3): 26–45.

