## Supplemental_Figure_1 for "Evaluating the CellSearch CMMC Assay for Non-Invasive Longitudinal MRD Monitoring"

* Contributed equally to this work

### Corresponding author

**Supplemental Figure 1.**

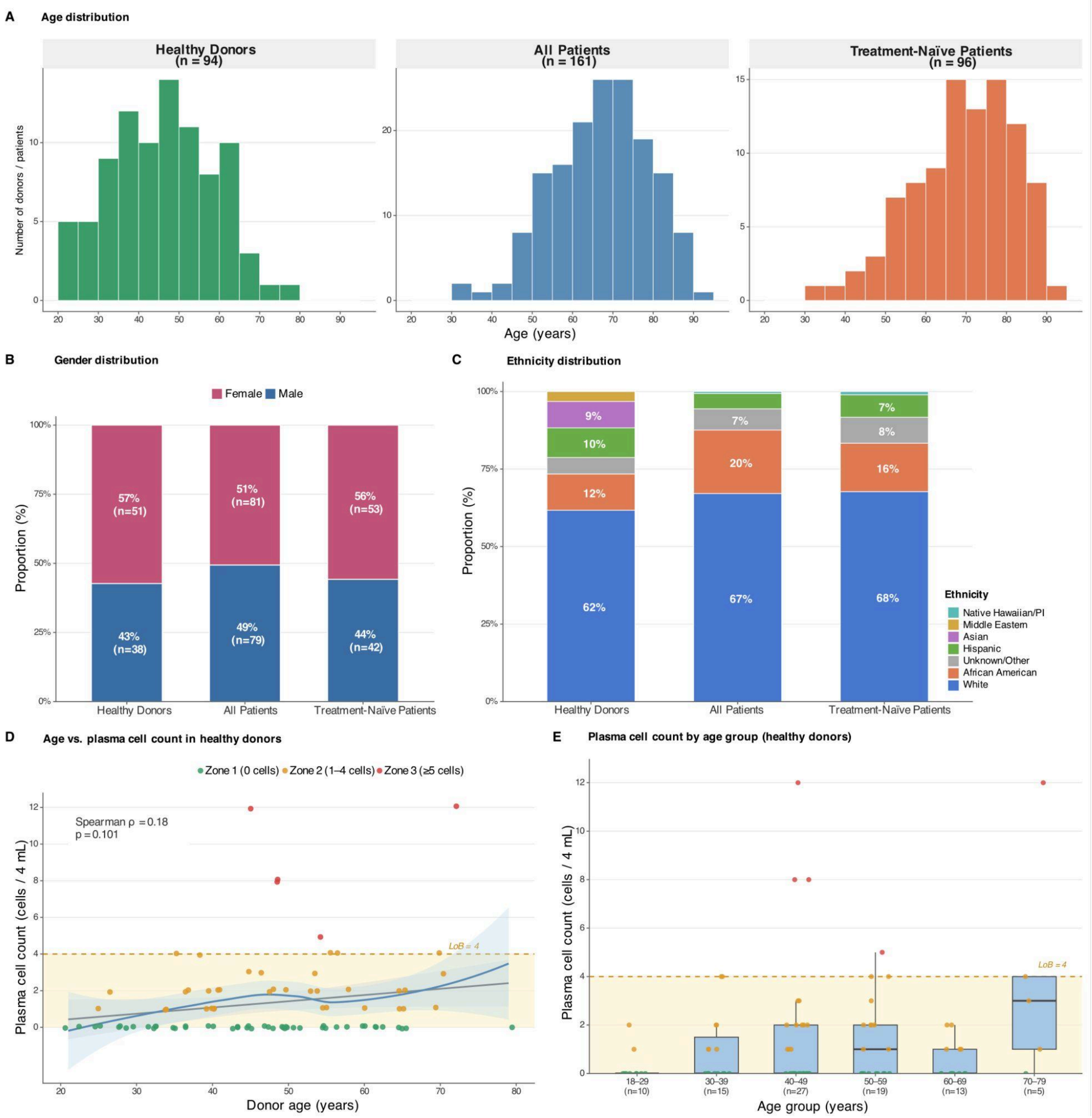


**Supplemental Figure 1. Cohort demographics and plasma cell background in healthy donors.**

**(A)** Age distribution of healthy donors (n = 94), all multiple myeloma patients (n = 161), and treatment-naïve patients (n = 96), shown as histograms with 5-year bins.

**(B)** Gender distribution across cohorts, expressed as proportions with absolute counts.

**(C)** Ethnicity distribution across cohorts; labels are shown for groups comprising ≥ 6% of the cohort. Granular self-reported ethnicity categories were collapsed into seven broad groups for display.

**(D)** Correlation between donor age and plasma cell count in healthy donors. Individual points are colored by the reporting zone (*Zone 1*: 0 cells; *Zone 2*: 1–4 cells; *Zone 3*: $\geq$ 5 cells). The grey band and line denote a linear regression fit with 95% confidence interval; the blue curve is a LOESS smoother (span = 0.9). The yellow shaded region indicates counts at or below the limit of blank (LoB = 4 cells / 4 mL blood). Spearman $\rho = 0.18$, $p = 0.10$.

**(E)** Plasma cell counts stratified by age decade in healthy donors. Boxes indicate median and interquartile range, individual points are colored by the reporting zone. A Kruskal–Wallis test showed no significant difference in plasma cell counts across age groups ($H(5) = 8.61$, $p = 0.13$).
