## Supplemental_Table_1 for "Evaluating the CellSearch CMMC Assay for Non-Invasive Longitudinal MRD Monitoring"

| Sample_ID | Group | Age | Gender | Ethnicity | Plasma_Count_Mean | CMMC_Assay_Replicates_Number | Raw_Plasma_Count_Data | WBC (x10^3/uL) | Treatment_Naive |
|---|---|---|---|---|---|---|---|---|---|
| ABS311817-01 | Patient | 31–35 | Female | African American/Non-Hispanic | 2 | 1 | 2 | 7.9 | no |
| ABS311817-02 | Patient | 61–65 | Male | African American/Non-Hispanic | 0 | 1 | 0 | 3.5 | no |
| ABS311817-03 | Patient | 66–70 | Male | White/Non-Hispanic | 36.5 | 2 | 37/36 | MISSING | no |
| ABS311817-04 | Patient | 51–55 | Male | African American/Non-Hispanic | 2.5 | 2 | 3/2 | MISSING | no |
| ABS311817-05 | Patient | 71–75 | Female | White/Non-Hispanic | 0 | 1 | 0 | MISSING | no |
| ABS311817-06 | Patient | 51–55 | Female | African American/Non-Hispanic | 233.5 | 2 | 233/234 | MISSING | no |
| ABS311817-07 | Patient | 61–65 | Male | White/Non-Hispanic | 0 | 1 | 0 | MISSING | no |
| ABS311817-08 | Patient | 51–55 | Male | White/Non-Hispanic | 7.5 | 2 | 7/8 | 40.4 | no |
| ABS311817-09 | Patient | 61–65 | Female | White/Non-Hispanic | 522 | 2 | 557/487 | MISSING | no |
| ABS311817-10 | Patient | 61–65 | Female | White/Non-Hispanic | 0.5 | 2 | 1/0 | 7.6 | no |
| ABS311817-11 | Patient | 56–60 | Male | African American/Non-Hispanic | 1.5 | 2 | 2/1 | 8.5 | no |
| ABS311817-12 | Patient | 56–60 | Male | African American/Non-Hispanic | 75.5 | 2 | 72/79 | 5 | no |
| ABS311817-13 | Patient | 56–60 | Male | White/Non-Hispanic | 0 | 2 | 0/0 | 4.4 | no |
| ABS311817-14 | Patient | 71–75 | Female | Unknown/Non-Hispanic | 6 | 2 | 5/7 | 5.3 | no |
| ABS311817-15 | Patient | 51–55 | Male | African American/Non-Hispanic | 119.5 | 2 | 119/120 | 5.8 | no |
| ABS311817-16 | Patient | 66–70 | Male | White/Non-Hispanic | 0 | 2 | 0/0 | 2.8 | no |
| ABS311817-17 | Patient | 61–65 | Male | African American/Non-Hispanic | 0 | 2 | 0/0 | 5.7 | no |
| ABS311817-18 | Patient | 61–65 | Male | White/Non-Hispanic | 17 | 2 | 17/17 | 6.7 | no |
| ABS311817-19 | Patient | 66–70 | Male | African American/Non-Hispanic | 16 | 2 | 16/16 | 5.2 | no |
| ABS311817-20 | Patient | 66–70 | Female | Unknown/Non-Hispanic | 0 | 2 | 0/0 | 7 | no |
| ABS311817-21 | Patient | 61–65 | Male | White/Non-Hispanic | 0 | 2 | 0/0 | 3.2 | no |
| ABS311817-22 | Patient | 76–80 | Male | White/Non-Hispanic | 376 | 2 | 375/377 | 5.9 | no |
| ABS311817-23 | Patient | 46–50 | Male | African American/Non-Hispanic | 1 | 2 | 1/1 | 4.4 | no |
| ABS311817-24 | Patient | 66–70 | Female | White/Non-Hispanic | 6 | 1 | 6 | 5.4 | no |
| ABS311817-26 | Patient | 71–75 | Male | White/Non-Hispanic | 1 | 1 | 1 | 9 | no |
| ABS311817-29 | Patient | 61–65 | Male | White/Non-Hispanic | 0.5 | 2 | 0/1 | 2.6 | no |
| ABS313965-01 | Patient | 41–45 | Female | White/Non-Hispanic | 154.5 | 2 | 149/160 | 6.5 | treatment_naive |
| ABS313965-02 | Patient | 51–55 | Female | African American/Non-Hispanic | 6 | 2 | 5/7 | 2.5 | treatment_naive |
| ABS313965-03 | Patient | 51–55 | Female | African American/Non-Hispanic | 24.5 | 2 | 24/25 | 4.9 | treatment_naive |
| ABS313063-01 | Patient | 61–65 | Female | African American/Non-Hispanic | 56 | 1 | 56 | 7 | no |
| ABS313063-02 | Patient | 51–55 | Male | White/Non-Hispanic | 0 | 1 | 0 | 3.8 | no |
| ABS313063-03 | Patient | 71–75 | Female | White/Non-Hispanic | 0 | 1 | 0 | 4.3 | no |

| Sample_ID | Group | Age | Gender | Ethnicity | Plasma_Count_Mean | CMMC_Assay_Replicates_Number | Raw_Plasma_Count_Data | WBC (x10^3/uL) | Treatment_Naive |
|---|---|---|---|---|---|---|---|---|---|
| ABS313063-04 | Patient | 46–50 | Female | African American/Non-Hispanic | 19 | 1 | 19 | 3 | no |
| ABS313063-05 | Patient | 61–65 | Female | African American/Non-Hispanic | 0 | 1 | 0 | 4.9 | no |
| ABS313063-06 | Patient | 81–85 | Male | White/Non-Hispanic | 1 | 1 | 1 | 5.8 | no |
| ABS313063-07 | Patient | 71–75 | Male | White/Non-Hispanic | 0 | 1 | 0 | 4.4 | no |
| ABS313063-08 | Patient | 66–70 | Female | White/Non-Hispanic | 0 | 1 | 0 | 3.1 | no |
| ABS313063-09 | Patient | 56–60 | Male | White/Non-Hispanic | 0 | 1 | 0 | 1.9 | no |
| ABS313063-10 | Patient | 71–75 | Male | African American/Non-Hispanic | 3 | 1 | 3 | 4.5 | no |
| ABS313063-11 | Patient | 66–70 | Female | White/Non-Hispanic | 0 | 1 | 0 | 4.3 | no |
| ABS313063-12 | Patient | 51–55 | Female | African American/Non-Hispanic | 743 | 1 | 743 | 3.4 | no |
| ABS313063-13 | Patient | 61–65 | Male | African American/Non-Hispanic | 0 | 1 | 0 | 4 | no |
| ABS313063-14 | Patient | 71–75 | Male | White/Non-Hispanic | 0.25 | 4 | 0/0/0/1 | 0.5 | no |
| ABS313063-15 | Patient | 71–75 | Female | White/Non-Hispanic | 0 | 4 | 0/0/0/0 | 8.5 | no |
| ABS313063-16 | Patient | 76–80 | Male | White/Non-Hispanic | 0 | 4 | 0/0/0/0 | 4.7 | no |
| ABS313063-17 | Patient | 56–60 | Female | White/Non-Hispanic | 0 | 4 | 0/0/0/0 | 6 | no |
| ABS313063-18 | Patient | 66–70 | Male | White/Non-Hispanic | 17.6 | 3 | 14/19/20 | 4.5 | no |
| ABS313063-19 | Patient | 71–75 | Male | White/Non-Hispanic | 0 | 4 | 0/0/0/0 | 6.9 | no |
| ABS313063-20 | Patient | 66–70 | Female | White/Non-Hispanic | 0 | 1 | 0 | 3.3 | no |
| ABS313063-21 | Patient | 61–65 | Male | White/Non-Hispanic | 84 | 1 | 84 | 8 | no |
| ABS313063-22 | Patient | 56–60 | Male | White/Non-Hispanic | 4 | 4 | 4/4/4/4 | 10.7 | no |
| ABS313063-23 | Patient | 56–60 | Female | White/Non-Hispanic | 515 | 1 | 515 | 11.2 | no |
| ABS313063-24 | Patient | 56–60 | Female | White/Non-Hispanic | 0 | 1 | 0 | 2.5 | no |
| ABS313063-25 | Patient | 46–50 | Female | White/Non-Hispanic | 0 | 1 | 0 | 7.7 | no |
| ABS313063-26 | Patient | 66–70 | Female | African American/Non-Hispanic | 260 | 1 | 260 | MISSING | no |
| ABS313063-27 | Patient | 51–55 | Male | White/Non-Hispanic | 0.5 | 2 | 1/0 | 2.8 | no |
| ABS313063-28 | Patient | 66–70 | Female | White/Non-Hispanic | 230.5 | 2 | 219/242 | 6.7 | no |
| ABS313063-29 | Patient | 51–55 | Female | African American/Non-Hispanic | 0 | 2 | 0/0 | 5 | no |
| ABS313063-30 | Patient | 46–50 | Male | Unknown | 1 | 2 | 1/1 | 11.7 | no |
| 202179211 | Patient | 71–75 | Female | Unknown | 5 | 2 | 4/6 | 10.2 | treatment_naive |
| 202179212 | Patient | 66–70 | Female | White/Non-Hispanic | 0 | 2 | 0/0 | 5.6 | treatment_naive |
| 202179214 | Patient | 61–65 | Female | White/Non-Hispanic | 5 | 2 | 4/6 | 4.9 | treatment_naive |
| 202179216 | Patient | 76–80 | Male | White/Non-Hispanic | 6 | 2 | 3/9 | MISSING | no |
| 202179233 | Patient | 61–65 | Male | White/Non-Hispanic | 146 | 2 | 148/144 | 4.1 | treatment_naive |

| Sample_ID | Group | Age | Gender | Ethnicity | Plasma_Count_Mean | CMMC_Assay_Replicates_Number | Raw_Plasma_Count_Data | WBC (x10^3/uL) | Treatment_Naive |
|---|---|---|---|---|---|---|---|---|---|
| 202179234 | Patient | 66–70 | Female | White/Non-Hispanic | 1 | 2 | 1/1 | 3.8 | treatment_naive |
| 202179235 | Patient | 86–90 | Female | White/Non-Hispanic | 3 | 2 | 3/3 | 5.9 | treatment_naive |
| 202179198 | Patient | 36–40 | Female | African American | 4 | 2 | 6/2 | 4 | treatment_naive |
| 202179208 | Patient | 76–80 | Female | White/Non-Hispanic | 19.5 | 2 | 21/18 | 4 | treatment_naive |
| 202179213 | Patient | 61–65 | Male | White/Non-Hispanic | 4 | 2 | 4/4 | 6.5 | treatment_naive |
| 202179237 | Patient | 46–50 | Female | Unknown | 3.5 | 2 | 4/3 | 5.4 | treatment_naive |
| 202179251 | Patient | 71–75 | Female | African American | 1213.5 | 2 | 1198/1229 | 5.4 | treatment_naive |
| 202179253 | Patient | 66–70 | Female | Unknown | 16.5 | 2 | 18/15 | 8.9 | treatment_naive |
| 202179209 | Patient | 51–55 | Female | White/Unknown | 2 | 2 | 2/2 | 5.7 | treatment_naive |
| 202179210 | Patient | 51–55 | Female | White/Non-Hispanic | 0 | 2 | 0/0 | 5.6 | treatment_naive |
| 202179236 | Patient | 71–75 | Female | White/Unknown | 0.5 | 2 | 0/1 | 4.4 | treatment_naive |
| 202179238 | Patient | 66–70 | Female | White/Non-Hispanic | 0.5 | 2 | 0/1 | 8.2 | treatment_naive |
| 202179200 | Patient | 46–50 | Female | African American/Non-Hispanic | 29.5 | 2 | 35/24 | 9.3 | treatment_naive |
| 202179201 | Patient | 51–55 | Female | White/Non-Hispanic | 1401 | 2 | 1300/1502 | 5.2 | treatment_naive |
| 202179215 | Patient | 56–60 | Male | African American (Native American | 6 | 2 | 8/4 | 3.1 | treatment_naive |
| 202179245 | Patient | 71–75 | Male | White/Non-Hispanic | 1.5 | 2 | 2/1 | 6.2 | treatment_naive |
| 202179244 | Patient | 71–75 | Male | White/Non-Hispanic | 3.5 | 2 | 0/7 | 2.9 | treatment_naive |
| 202179232 | Patient | 76–80 | Female | White/Non-Hispanic | 0 | 2 | 0/0 | 5.1 | treatment_naive |
| 202179248 | Patient | 81–85 | Male | White/Hispanic | 312.5 | 2 | 310/315 | 4.9 | treatment_naive |
| 202179239 | Patient | 81–85 | Female | White/Hispanic | 107.5 | 2 | 107/108 | 5.5 | no |
| 202179246 | Patient | 86–90 | Male | White/Unknown | 8 | 2 | 7/9 | 5.1 | treatment_naive |
| 202179249 | Patient | 81–85 | Female | White/Non-Hispanic | 2 | 2 | 2/2 | 4.1 | treatment_naive |
| 202179205 | Patient | 61–65 | Male | African American | 235 | 2 | 238/232 | 6.8 | treatment_naive |
| 202179206 | Patient | 56–60 | Male | White/Non-Hispanic | 134 | 2 | 125/143 | 5.8 | treatment_naive |
| 202179207 | Patient | 66–70 | Male | White/Non-Hispanic | 0.5 | 2 | 0/1 | 4.5 | treatment_naive |
| 202179252 | Patient | 71–75 | Male | African American | 35 | 2 | 33/37 | 3.6 | treatment_naive |
| 202179258 | Patient | 71–75 | Female | White/Non-Hispanic | 0 | 2 | 0/0 | 5.7 | treatment_naive |
| 202179255 | Patient | 66–70 | Male | White/Non-Hispanic | 23 | 2 | 26/20 | 4.6 | treatment_naive |
| 202179256 | Patient | 81–85 | Male | White/Non-Hispanic | 4 | 2 | 4/4 | 9.9 | treatment_naive |
| 202184805 | Patient | 76–80 | Male | White/Non-Hispanic | 1121 | 2 | 1158/1084 | 2.9 | treatment_naive |
| 202179202 | Patient | 86–90 | Female | White/Non-Hispanic | 1 | 2 | 1/1 | 7.8 | treatment_naive |
| 202179240 | Patient | 76–80 | Female | White/Non-Hispanic | 0.5 | 2 | 0/1 | 4.3 | treatment_naive |

| Sample_ID | Group | Age | Gender | Ethnicity | Plasma_Count_Mean | CMMC_Assay_Replicates_Number | Raw_Plasma_Count_Data | WBC (x10^3/uL) | Treatment_Naive |
|---|---|---|---|---|---|---|---|---|---|
| 202179254 | Patient | 86–90 | Male | White/Non-Hispanic | 26 | 2 | 26/26 | 7.5 | treatment_naive |
| 202179203 | Patient | 51–55 | Female | African American/Non-Hispanic | 39.5 | 2 | 39/40 | 3.6 | treatment_naive |
| 202179204 | Patient | 81–85 | Male | White/Unknown | 5 | 2 | 5/5 | 5.2 | treatment_naive |
| 202179241 | Patient | 76–80 | Male | White/Non-Hispanic | 2 | 2 | 2/2 | 4.8 | treatment_naive |
| 202179247 | Patient | 61–65 | Male | White/Non-Hispanic | 6786.5 | 2 | 6852/6721 | 4.9 | treatment_naive |
| 202184661 | Patient | 81–85 | Female | White/Non-Hispanic | 1505 | 2 | 1455/1555 | 5.8 | treatment_naive |
| 202184801 | Patient | 46–50 | Male | White/Unknown | 239.5 | 2 | 235/244 | MISSING | no |
| 202184662 | Patient | 66–70 | Male | Unknown/Hispanic | 2 | 2 | 2/2 | 3.2 | treatment_naive |
| 202184660 | Patient | 76–80 | Male | White/Non-Hispanic | 15 | 2 | 18/12 | 4.3 | treatment_naive |
| 202179250 | Patient | 61–65 | Female | White/Non-Hispanic | 4 | 2 | 4/4 | 5.8 | treatment_naive |
| 202184802 | Patient | 51–55 | Male | White/Unknown | 2 | 2 | 2/2 | 5.7 | treatment_naive |
| 202184803 | Patient | 41–45 | Male | White/Non-Hispanic | 1.5 | 2 | 1/2 | 5.7 | treatment_naive |
| 202184804 | Patient | 56–60 | Male | White/Non-Hispanic | 2 | 2 | 2/2 | 5.7 | treatment_naive |
| 202188453 | Patient | 76–80 | Female | White/Non-Hispanic | 116.5 | 2 | 110/123 | 4 | treatment_naive |
| 202179197 | Patient | 46–50 | Female | African American | 19.5 | 2 | 18/21 | 3.5 | treatment_naive |
| 202188445 | Patient | 76–80 | Female | White/Non-Hispanic | 4 | 2 | 4/4 | MISSING | no |
| 202188455 | Patient | 71–75 | Male | White/Non-Hispanic | 3649.5 | 2 | 3796/3503 | MISSING | no |
| 202184663 | Patient | 71–75 | Male | White/Non-Hispanic | 0 | 2 | 0/0 | MISSING | no |
| 202188459 | Patient | 76–80 | Female | White/Hispanic | 1438 | 2 | 1458/1418 | 3.9 | treatment_naive |
| 202185276 | Patient | 81–85 | Female | Unknown/Hispanic | 9.5 | 2 | 9/10 | MISSING | no |
| 202188449 | Patient | 81–85 | Male | White/Non-Hispanic | 6 | 2 | 5/7 | 3.3 | treatment_naive |
| 202188456 | Patient | 76–80 | Female | White/Non-Hispanic | 6 | 2 | 6/6 | 4.7 | treatment_naive |
| 202184664 | Patient | 66–70 | Female | Unknown/Hispanic | 9.5 | 2 | 9/10 | 6.2 | treatment_naive |
| 202188457 | Patient | 56–60 | Female | Native Hawaiian/Non-Hispanic | 4.5 | 2 | 5/4 | 5.7 | treatment_naive |
| 202188633 | Patient | Unknown | Unknown | Unknown | 10.5 | 2 | 9/12 | 5.2 | treatment_naive |
| 202188634 | Patient | 86–90 | Male | White/Non-Hispanic | 80.5 | 2 | 84/77 | 5.3 | treatment_naive |
| 202188636 | Patient | 71–75 | Male | White/Non-Hispanic | 2.5 | 2 | 3/2 | MISSING | no |
| 202188637 | Patient | 81–85 | Female | Unknown/Hispanic | 37.5 | 2 | 35/40 | 3 | treatment_naive |
| 202188635 | Patient | 76–80 | Male | African American/Non-Hispanic | 11 | 2 | 12/10 | 3.4 | treatment_naive |
| 202188450 | Patient | 66–70 | Male | White/Non-Hispanic | 40.5 | 2 | 39/42 | 1.9 | treatment_naive |
| 202188452 | Patient | 56–60 | Male | White/Non-Hispanic | 0.5 | 2 | 1/0 | 5.2 | treatment_naive |
| 202188454 | Patient | 61–65 | Male | White/Non-Hispanic | 558 | 2 | 575/541 | 6.3 | treatment_naive |

| Sample_ID | Group | Age | Gender | Ethnicity | Plasma_Count_Mean | CMMC_Assay_Replicates_Number | Raw_Plasma_Count_Data | WBC (x10^3/uL) | Treatment_Naive |
|---|---|---|---|---|---|---|---|---|---|
| 202188639 | Patient | 71–75 | Male | White/Non-Hispanic | 0 | 2 | 0/0 | 3.8 | treatment_naive |
| 202188446 | Patient | 61–65 | Female | Unknown | 1 | 2 | 2/0 | 3 | treatment_naive |
| 202188447 | Patient | 66–70 | Female | White/Non-Hispanic | 21.5 | 2 | 21/22 | 6.7 | treatment_naive |
| 202188451 | Patient | 76–80 | Female | White/Non-Hispanic | 1.5 | 2 | 2/1 | 5.9 | treatment_naive |
| 202188458 | Patient | 66–70 | Female | Unknown/Hispanic | 766.5 | 2 | 758/775 | 12.5 | treatment_naive |
| 202188638 | Patient | 76–80 | Male | African American/Non-Hispanic | 6 | 2 | 6/6 | 2.8 | treatment_naive |
| 202188640 | Patient | 71–75 | Female | Unknown | 119.5 | 2 | 121/118 | 5.6 | treatment_naive |
| 202185277 | Patient | 86–90 | Female | White/Non-Hispanic | 1.5 | 2 | 2/1 | 5.3 | treatment_naive |
| 202188645 | Patient | 66–70 | Male | White/Non-Hispanic | 0.5 | 2 | 0/1 | 5.7 | treatment_naive |
| 202188448 | Patient | 91–95 | Male | White/Non-Hispanic | 0.5 | 2 | 1/0 | 3.6 | treatment_naive |
| 202179261 | Patient | 71–75 | Female | White/Non-Hispanic | 2 | 2 | 2/2 | 6.4 | treatment_naive |
| 202185279 | Patient | 76–80 | Male | White/Non-Hispanic | 2.5 | 2 | 2/3 | 6.1 | treatment_naive |
| 202196057 | Patient | 56–60 | Female | Unknown/Non-Hispanic | 94 | 2 | 101/87 | 3.8 | treatment_naive |
| 202196060 | Patient | 71–75 | Female | White/Non-Hispanic | 3 | 2 | 5/1 | MISSING | no |
| 202185278 | Patient | 76–80 | Female | White/Non-Hispanic | 0 | 2 | 0/0 | 3.1 | treatment_naive |
| 202196058 | Patient | 81–85 | Male | White/Non-Hispanic | 5.5 | 2 | 6/5 | 4.6 | treatment_naive |
| 202196059 | Patient | 66–70 | Female | Unknown/Non-Hispanic | 48 | 2 | 47/49 | 10 | treatment_naive |
| 202196053 | Patient | 71–75 | Female | White/Non-Hispanic | 34.5 | 2 | 33/36 | 5.1 | treatment_naive |
| 202196055 | Patient | 66–70 | Female | White/Non-Hispanic | 9.5 | 2 | 9/10 | 4.3 | treatment_naive |
| 202196056 | Patient | 66–70 | Female | African American/Non-Hispanic | 425.5 | 2 | 420/431 | 5.2 | treatment_naive |
| 202188642 | Patient | 86–90 | Male | Unknown/Hispanic | 13 | 2 | 14/12 | 8.2 | treatment_naive |
| 202185280 | Patient | 81–85 | Female | White/Non-Hispanic | 78 | 2 | 92/64 | 5 | treatment_naive |
| 202188641 | Patient | 56–60 | Female | Unknown/Hispanic | 0.5 | 2 | 1/0 | 2.3 | treatment_naive |
| 202196044 | Patient | 81–85 | Female | White/Non-Hispanic | 1 | 2 | 1/1 | 3.7 | treatment_naive |
| 202179257 | Patient | 71–75 | Female | White/Non-Hispanic | 47.5 | 2 | 50/45 | 4.7 | treatment_naive |
| 202188643 | Patient | 76–80 | Male | African American/Non-Hispanic | 99.5 | 2 | 99/100 | 4.3 | treatment_naive |
| 202188644 | Patient | 31–35 | Female | Unknown/Hispanic | 16 | 2 | 18/14 | 7.1 | treatment_naive |
| 202188207 | Patient | 81–85 | Male | White/Non-Hispanic | 7.5 | 2 | 8/7 | 6.5 | treatment_naive |
| 202188210 | Patient | 81–85 | Female | White/Non-Hispanic | 0.5 | 2 | 0/1 | 3.3 | treatment_naive |
| 202188204 | Patient | 61–65 | Male | African American/Unknown | 110 | 2 | 107/113 | 7 | treatment_naive |
| 202188208 | Patient | 56–60 | Male | White/Non-Hispanic | 414.5 | 2 | 422/407 | 4.8 | treatment_naive |
| 202188209 | Patient | 86–90 | Male | White/Non-Hispanic | 1.5 | 2 | 2/1 | 1.8 | treatment_naive |

| Sample_ID | Group | Age | Gender | Ethnicity | Plasma_Count_Mean | CMMC_Assay_Replicates_Number | Raw_Plasma_Count_Data | WBC (x10^3/uL) | Treatment_Naive |
|---|---|---|---|---|---|---|---|---|---|
| 202193828 | Patient | 71–75 | Male | White/Non-Hispanic | 2 | 2 | 4/0 | 6.7 | treatment_naive |
| B21-17 | Healthy | 46–50 | Male | White/Non-Hispanic | 0 | 1 | 0 | MISSING | Healthy |
| B21-185 | Healthy | 66–70 | Female | White/Non-Hispanic | 3 | 1 | 3 | MISSING | Healthy |
| B21-186 | Healthy | 46–50 | Male | Middle Eastern | 2 | 1 | 2 | MISSING | Healthy |
| B21-187 | Healthy | 66–70 | Female | White/Non-Hispanic | 1 | 1 | 1 | MISSING | Healthy |
| B21-188 | Healthy | 61–65 | Male | White/Non-Hispanic | 0 | 1 | 0 | MISSING | Healthy |
| B21-189 | Healthy | 31–35 | Male | White/Non-Hispanic | 0 | 1 | 0 | MISSING | Healthy |
| B21-190 | Healthy | 46–50 | Male | White/Non-Hispanic | 2 | 1 | 2 | MISSING | Healthy |
| B21-191 | Healthy | 61–65 | Female | White/Non-Hispanic | 0 | 1 | 0 | MISSING | Healthy |
| B21-192 | Healthy | 56–60 | Male | Asian | 4 | 1 | 4 | MISSING | Healthy |
| B21-200 | Healthy | 36–40 | Female | White/Non-Hispanic | 0 | 1 | 0 | MISSING | Healthy |
| B21-202 | Healthy | 56–60 | Female | White/Non-Hispanic | 0 | 1 | 0 | MISSING | Healthy |
| B21-203 | Healthy | 51–55 | Male | African American | 0 | 1 | 0 | MISSING | Healthy |
| B21-204 | Healthy | 36–40 | Male | White/Non-Hispanic | 0 | 1 | 0 | MISSING | Healthy |
| B21-205 | Healthy | 66–70 | Female | White/Non-Hispanic | 4 | 1 | 4 | MISSING | Healthy |
| B21-206 | Healthy | 31–35 | Male | Asian | 0 | 1 | 0 | MISSING | Healthy |
| B21-208 | Healthy | 56–60 | Female | White/Non-Hispanic | 2 | 1 | 2 | MISSING | Healthy |
| B21-21 | Healthy | 41–45 | Male | Middle Eastern | 0 | 1 | 0 | MISSING | Healthy |
| B21-210 | Healthy | 46–50 | Female | African American | 0 | 1 | 0 | MISSING | Healthy |
| B21-217 | Healthy | 31–35 | Female | White/Non-Hispanic | 0 | 1 | 0 | MISSING | Healthy |
| B21-218 | Healthy | 31–35 | Male | Hispanic | 1 | 1 | 1 | MISSING | Healthy |
| B21-219 | Healthy | 56–60 | Female | White/Non-Hispanic | 1 | 1 | 1 | MISSING | Healthy |
| B21-22 | Healthy | 56–60 | Female | Hispanic | 0 | 1 | 0 | MISSING | Healthy |
| B21-220 | Healthy | 46–50 | Male | White/Non-Hispanic | 0 | 1 | 0 | MISSING | Healthy |
| B21-221 | Healthy | 56–60 | Female | White/Non-Hispanic | 0 | 1 | 0 | MISSING | Healthy |
| B21-222 | Healthy | 41–45 | Female | Middle Eastern | 0 | 1 | 0 | MISSING | Healthy |
| B21-223 | Healthy | 61–65 | Male | White/Non-Hispanic | 0 | 1 | 0 | MISSING | Healthy |
| B21-23 | Healthy | 41–45 | Female | White/Non-Hispanic | 0 | 1 | 0 | MISSING | Healthy |
| B21-235 | Healthy | 36–40 | Male | White/Non-Hispanic | 0 | 1 | 0 | MISSING | Healthy |
| B21-237 | Healthy | 31–35 | Male | African American | 4 | 1 | 4 | MISSING | Healthy |
| B21-239 | Healthy | 46–50 | Female | White/Non-Hispanic | 3 | 1 | 3 | MISSING | Healthy |
| B21-24 | Healthy | 61–65 | Female | White/Non-Hispanic | 0 | 1 | 0 | MISSING | Healthy |

| Sample_ID | Group | Age | Gender | Ethnicity | Plasma_Count_Mean | CMMC_Assay_Replicates_Number | Raw_Plasma_Count_Data | WBC (x10^3/uL) | Treatment_Naive |
|---|---|---|---|---|---|---|---|---|---|
| B21-240 | Healthy | 41–45 | Male | African American | 12 | 1 | 12 | MISSING | Healthy |
| B21-241 | Healthy | 61–65 | Female | White/Non-Hispanic | 0 | 1 | 0 | MISSING | Healthy |
| B21-246 | Healthy | 51–55 | Female | White/Non-Hispanic | 0 | 1 | 0 | MISSING | Healthy |
| B21-247 | Healthy | 46–50 | Female | Hispanic | 0 | 1 | 0 | MISSING | Healthy |
| B21-25 | Healthy | 46–50 | Female | African American | 0 | 1 | 0 | MISSING | Healthy |
| B21-27 | Healthy | 71–75 | Male | African American | 12 | 1 | 12 | MISSING | Healthy |
| B21-30 | Healthy | 61–65 | Female | White/Non-Hispanic | 2 | 1 | 2 | MISSING | Healthy |
| B21-31 | Healthy | 36–40 | Male | White/Non-Hispanic | 1 | 1 | 1 | MISSING | Healthy |
| B21-32 | Healthy | 36–40 | Female | White/Non-Hispanic | 1 | 1 | 1 | MISSING | Healthy |
| B21-33 | Healthy | 36–40 | Male | White/Non-Hispanic | 4 | 1 | 4 | MISSING | Healthy |
| B21-34 | Healthy | 51–55 | Male | White/Non-Hispanic | 2 | 1 | 2 | MISSING | Healthy |
| B21-36 | Healthy | 36–40 | Male | Asian | 2 | 1 | 2 | MISSING | Healthy |
| B21-38 | Healthy | 51–55 | Male | White/Non-Hispanic | 1 | 1 | 1 | MISSING | Healthy |
| B21-40 | Healthy | 36–40 | Male | Asian | 1 | 1 | 1 | MISSING | Healthy |
| B21-41 | Healthy | 46–50 | Female | Hispanic | 0 | 1 | 0 | MISSING | Healthy |
| B21-42 | Healthy | 76–80 | Female | Hispanic | 0 | 1 | 0 | MISSING | Healthy |
| B21-43 | Healthy | 41–45 | Male | African American | 3 | 1 | 3 | MISSING | Healthy |
| B21-44 | Healthy | 61–65 | Female | White/Non-Hispanic | 1 | 1 | 1 | MISSING | Healthy |
| B21-45 | Healthy | 41–45 | Female | White/Non-Hispanic | 0 | 1 | 0 | MISSING | Healthy |
| B22-288 | Healthy | 36–40 | Female | White/Non-Hispanic | 1 | 1 | 1 | MISSING | Healthy |
| B22-289 | Healthy | 56–60 | Female | White/Non-Hispanic | 0 | 1 | 0 | MISSING | Healthy |
| B22-294 | Healthy | 26–30 | Female | Hispanic | 0 | 1 | 0 | MISSING | Healthy |
| B22-309 | Healthy | 51–55 | Male | White/Non-Hispanic | 0 | 1 | 0 | MISSING | Healthy |
| B22-310 | Healthy | 21–25 | Female | White/Non-Hispanic | 0 | 1 | 0 | MISSING | Healthy |
| B22-328 | Healthy | 21–25 | Male | Hispanic | 0 | 1 | 0 | MISSING | Healthy |
| B22-333 | Healthy | 46–50 | Male | White/Non-Hispanic | 0 | 1 | 0 | 4 | Healthy |
| B22-337 | Healthy | 26–30 | Male | African American | 0 | 1 | 0 | MISSING | Healthy |
| B22-350 | Healthy | 51–55 | Female | White/Non-Hispanic | 5 | 1 | 5 | MISSING | Healthy |
| B22-351 | Healthy | 61–65 | Female | White/Non-Hispanic | 0 | 1 | 0 | MISSING | Healthy |
| B22-352 | Healthy | 46–50 | Female | White/Non-Hispanic | 2 | 1 | 2 | MISSING | Healthy |
| B22-362 | Healthy | 21–25 | Female | White/Non-Hispanic | 0 | 1 | 0 | MISSING | Healthy |
| B22-363 | Healthy | 41–45 | Female | White/Non-Hispanic | 0 | 1 | 0 | MISSING | Healthy |

| Sample_ID | Group | Age | Gender | Ethnicity | Plasma_Count_Mean | CMMC_Assay_Replicates_Number | Raw_Plasma_Count_Data | WBC (x10^3/uL) | Treatment_Naive |
|---|---|---|---|---|---|---|---|---|---|
| B22-367 | Healthy | Unknown | Unknown | Unknown | 0 | 1 | 0 | MISSING | Healthy |
| B22-368A | Healthy | 51–55 | Female | White/Non-Hispanic | 0 | 1 | 0 | MISSING | Healthy |
| B22-368B | Healthy | 51–55 | Female | White/Non-Hispanic | 1 | 1 | 1 | MISSING | Healthy |
| B22-369A | Healthy | 46–50 | Female | African American | 8 | 1 | 8 | MISSING | Healthy |
| B22-369B | Healthy | 46–50 | Female | African American | 8 | 1 | 8 | MISSING | Healthy |
| B22-370A | Healthy | 31–35 | Male | White/Non-Hispanic | 0 | 1 | 0 | MISSING | Healthy |
| B22-370B | Healthy | 31–35 | Male | White/Non-Hispanic | 0 | 1 | 0 | MISSING | Healthy |
| B22-385 | Healthy | Unknown | Unknown | Unknown | 0 | 1 | 0 | MISSING | Healthy |
| B22-391A | Healthy | 41–45 | Male | Asian | 2 | 1 | 2 | MISSING | Healthy |
| B22-391B | Healthy | 41–45 | Male | Asian | 2 | 1 | 2 | MISSING | Healthy |
| B22-392 | Healthy | Unknown | Unknown | Unknown | 2 | 1 | 2 | MISSING | Healthy |
| B22-393 | Healthy | 51–55 | Female | White/Non-Hispanic | 3 | 1 | 3 | 7.8 | Healthy |
| B22-395 | Healthy | 61–65 | Female | White/Non-Hispanic | 1 | 1 | 1 | MISSING | Healthy |
| B22-396 | Healthy | Unknown | Unknown | Unknown | 0 | 1 | 0 | MISSING | Healthy |
| B22-405 | Healthy | Unknown | Unknown | Unknown | 0 | 1 | 0 | 4.9 | Healthy |
| B22-432 | Healthy | 51–55 | Female | Hispanic | 2 | 1 | 2 | 5.6 | Healthy |
| B22-433 | Healthy | 46–50 | Male | Hispanic | 0 | 1 | 0 | 6.1 | Healthy |
| B22-436 | Healthy | 31–35 | Female | White/Non-Hispanic | 1 | 1 | 1 | 4.7 | Healthy |
| B22-437 | Healthy | 36–40 | Female | White/Non-Hispanic | 0 | 1 | 0 | 5.5 | Healthy |
| B22-438 | Healthy | 21–25 | Female | White/Non-Hispanic | 0 | 1 | 0 | 5.4 | Healthy |
| B22-439 | Healthy | 26–30 | Female | White/Non-Hispanic | 2 | 1 | 2 | 6.6 | Healthy |
| B22-441 | Healthy | 61–65 | Female | White/Non-Hispanic | 2 | 1 | 2 | 6.3 | Healthy |
| B22-477 | Healthy | 51–55 | Female | African American | 0 | 1 | 0 | MISSING | Healthy |
| B23-016 | Healthy | 31–35 | Female | White/Non-Hispanic | 0 | 1 | 0 | 8.4 | Healthy |
| B23-020 | Healthy | 26–30 | Male | White/Non-Hispanic | 0 | 1 | 0 | 4.2 | Healthy |
| B23-063A | Healthy | 36–40 | Male | White/Non-Hispanic | 2 | 1 | 2 | 7.5 | Healthy |
| B23-063B | Healthy | 36–40 | Male | White/Non-Hispanic | 2 | 1 | 2 | 7.5 | Healthy |
| B23-094 | Healthy | 21–25 | Female | Asian | 1 | 1 | 1 | 4.99 | Healthy |
| B23-134 | Healthy | 41–45 | Male | Asian | 0 | 1 | 0 | 5.5 | Healthy |
| B23-163 | Healthy | 56–60 | Male | White/Non-Hispanic | 4 | 1 | 4 | 8 | Healthy |
| B23-164 | Healthy | 26–30 | Female | White/Non-Hispanic | 0 | 1 | 0 | 6.7 | Healthy |
